# Cognitive Trajectories and Clinical Outcomes in the North American Psychosis-Proneness Longitudinal Study (NAPLS2)

**DOI:** 10.64898/2026.11.24.26367563

**Authors:** Walid Yassin, James B. Green, Michael Cai, Danah Ansari, Kristen Woodbury, Daniel I. Shapiro, Kelly Allott, Siyan He, Xue-Jun Kong, Matcheri Keshavan, Jean Addington, Carrie E. Bearden, Kristin S. Cadenhead, Tyrone D. Cannon, Daniel H. Mathalon, Diana O. Perkins, Elaine F. Walker, Scott W. Woods, Paulo Fusar-Poli, William S. Stone

## Abstract

Cognitive impairments are well-documented in individuals at clinical high risk for psychosis (CHR), particularly among those who convert (i.e., transition) to psychosis. However, considerable heterogeneity in cognitive profiles across CHR populations limits the utility of cognition as a predictive or treatment-relevant marker. This study examined baseline and longitudinal (12 mo and 24 mo) neurocognitive functioning in CHR individuals from the North American Prodrome Longitudinal Study 2 (NAPLS 2) who either transitioned to psychosis or remitted, or who neither converted nor remitted over a 2-year follow-up period. A total of 689 CHR individuals and 265 healthy controls (HC) completed neurocognitive and clinical assessments. CHR participants were categorized based on clinical outcomes: converters (n=89), remitters (n=216), and CHR who neither converted nor remitted (n=384). Group differences at baseline and over time were assessed using ANCOVA and mixed-effects modeling, adjusting for demographic and site-related factors. At baseline, all CHR groups performed worse than HC across nearly all cognitive domains. Subsequent converters showed significantly lower baseline performance than remitters and those who remained CHR throughout the study in key domains, suggestive of state-like effects. All groups, including converters, showed different levels of improvements associated with practice effects. CHR in remission showed cognitive improvements, but not at levels associated with full recovery on most assessments. Our findings suggest that cognitive trajectories within clinical outcome are as heterogenous as those between, suggesting the importance of addressing such differences on a participant level.

## Introduction

Cognitive deficits constitute common features of schizophrenia (SCZ) that occur in all phases or stages of the disorder (1–5). Individuals with SCZ show significant cognitive deficits on average (6) that often reach 1.0 or more standard deviations below relevant general population means (7). There is considerable heterogeneity (2,8), however, including a minority of individuals (∼20%) who perform in the average range of cognitive performance (9–11) or higher (11,12), which reflects greater variability in SCZ spectrum conditions than is evident in healthy control (HC) subjects (2,11,13). Notably, it is unclear whether or to what extent average cognitive performances reflect declines from higher levels, as opposed to showing immunity to decline. Although individuals at clinical high risk for psychosis may show differences in high risk syndromes (e.g., some show a ‘genetic risk and deterioration’ syndrome or a ‘brief intermittent psychotic’ syndrome), most individuals ascertained for study meet diagnostic criteria for a clinical high risk for psychosis (CHR) syndrome involving attenuated psychosis (14–17). A recent meta-analysis assessing the largest CHR sample to date (n=5162) largely confirmed widespread cognitive deficits in CHR (18). Many, though not all, cognitive tests included in that study showed effect sizes in the small to medium range, compared to HC. A few tests showing effect sizes in the medium to large range involved working memory, processing speed, and learning, as shown, for example, in Seidman et al (4). Consistent with many of the individual studies comprising the meta-analyses, we and others have shown smaller, significant cognitive differences between CHR who developed psychosis compared to CHR who did not (11). Transition to psychosis was most evident in performance declines in tests of learning and memory, processing speed, attention/vigilance, and IQ. In a follow-up meta-analysis of these data using variability ratios (13), which compared variability in individuals with CHR and first episode psychosis (FEP) to variability in HC, CHR individuals showed higher variability ratios compared to HC in several cognitive domains, most notably learning and memory, premorbid IQ, processing speed, and executive functioning. Individuals with CHR who transitioned to psychosis showed greater variability ratios in executive functions compared to both those with CHR who did not transition and to individuals with FEP. Despite these relative differences between CHR and FEP; however, we also observed greater variability in FEP (n=3,925) compared to HC (n=1,285) in another recent meta-analysis (8).

Increased levels of cognitive variability in CHR and FEP highlight the greater cognitive heterogeneity that is associated with SCZ and similar conditions. Heterogeneity limits our understanding of at least two of the most important reasons to study cognition in SCZ, including 1) cognition’s role as a marker of clinical outcomes, and 2) cognition’s role(s) in facilitating/reducing adaptive functions such as succeeding in school, living independently, and obtaining and maintaining employment (19). Other sources of variability complicate our understanding of cognition in SCZ as well. Many cognitive variables are correlated significantly with each other, for example, though often not highly with clinical diagnoses or symptoms of psychosis. Moreover, cognitive performance is subject to influence from a host of internal (e.g., clinical comorbidities, genetic and epigenetic effects) and external factors (lack of appropriate cognitive tests and norms for use in low-and middle-income countries and for diverse subgroups in high- and in low-and middle-income countries, as well as practice effects, medical problems, effects of medications, effects of substance use and other adverse environmental effects).

Despite the heterogeneity, cognition in CHR has been validated as a significant feature associated with the development of illness, as shown, for example, by its inclusion in risk calculators for psychosis (20–22). Moreover, cognitive performances in CHR who subsequently transition to psychosis in the following 1-2 years are often lower at baseline than in CHR who do not transition to psychosis over the same period of time (18,23,24). Stratification of CHR into groups that do or do not transition to psychosis thus provides a useful, if not yet sufficient, strategy to reduce heterogeneity and increase the utility of cognitive assessment (11).

Prior to the ongoing AMP SCZ study (25), the North American Prodrome Longitudinal Study (NAPLS) was among the largest initiatives to investigate relationships between cognition and clinical outcomes. A case-control study using the NAPLS-1 cohort (CHR n=304) and accounting for effects of age, gender, parental education, site, and IQ showed widespread deficits in CHR compared with healthy controls (26). CHR who transitioned to psychosis showed larger deficits (*d*≈0.4) compared to CHR who did not transition, and highlighted domains (verbal learning, processing speed) showing the largest deficits and associations with future conversions. Baseline relationships of CHR converters with later clinical outcomes were also assessed with the NAPLS-2 (CHR n=689) cohort (4), which identified attention / working memory and declarative learning and memory as among the strongest differences versus HC subjects (*d*≈0.80), and also identified small (*d*=0.28 for attention/working memory) to small/medium (*d*=0.48) deficits versus CHR who did not develop psychosis over the course of the study. Lower verbal premorbid and declarative memory cognitive abilities were among the factors associated with reduced time to develop psychosis.

The current study extends this work by focusing on the NAPLS-2 longitudinal data, which were collected over a 2-year follow-up period. Longitudinal assessment adds a more dynamic perspective to understanding pathways to psychosis that takes advantage of additional changes, both in direction and magnitude, that lead to clinical outcomes. It may show, for example, that the onset of psychosis is not necessarily associated with additional deterioration of cognitive performance (27,28) or highlight associations with other outcomes, such as remission. In the current study, we will use the same clinical criteria for remission that were developed using NAPLS-3 and validated in NAPLS-2 (29), and will compare a remitter group in the same model with CHR groups who do and do not transition to psychosis, as well as the HC group.

## Methods

### Participants

The participant cohort included in this study is part of the NAPLS-2. Baseline and longitudinal neurocognitive data collected between 2009 and 2015 included 689 CHR participants and 265 HC. Baseline neurocognitive data were available for all participants; longitudinal neurocognitive data included 440 CHR participants and 198 HC. After follow-ups, the CHR participants were divided further into 66 converters (CHR-C), 181 remitters (CHR-R), and 193 non-converter, non-remitters (CHR-NCNR). Remission here is defined as scoring below the prodromal risk threshold (≤2) on all five positive symptom subscale items of the SOPS. The protocols for the study were approved by institutional review boards at each of the participating sites, with all participants providing written informed consent. Briefly, 12- to 35-year-old participants had to meet Criteria of Prodromal Syndromes (COPS), based on the Structured Interview for Prodromal Syndromes (SIPS) or criteria for DSM-IV schizotypal personality disorder if they were under 19. Exclusion criteria included a lifetime Axis I psychotic disorder, an IQ below 70, DSM-IV substance dependence in the past 6 months, or a central nervous system disorder that was likely to interfere with cognitive or clinical measures collected in the study. The HC participants did not meet criteria for a psychosis-proneness syndrome, current or past psychotic disorder, or cluster A personality disorder. Their first-degree relatives did not have a history of psychotic disorder or symptoms.

### Clinical Measures

In order to rule out psychosis and identify DSM-IV Axis I or cluster A, Axis II personality disorders, the Structured Clinical Interview (SCID) for DSM-IV (SCID-IV) was used (30). Psychosis symptomology was measured using the SOPS criteria (31). The SIPS Presence of Psychotic Symptoms criteria were used to determine the transition to psychosis.

### Neurocognitive Test Battery

The battery included measures of neurocognitive functioning that are often compromised in SCZ spectrum conditions, and some we did not expect to show commensurate changes, such as the Wide Range Achievement Test - 4th Edition (WRAT-4) Reading Task (32), which was used for premorbid IQ estimates. Current IQ estimates were obtained using a two-subtest version of the Wechsler Abbreviated Scale of Intelligence (WASI), with the Vocabulary (WASI Vocab) and Block design (WASI BD) subtests (33). Other cognitive domains were assessed using subtests from the Measurement and Treatment Research to Improve Cognition in Schizophrenia (MATRICS) Consensus Cognitive Battery (MCCB), including Hopkins Verbal Learning Test-Revised (HVLT-R) (34), and the Brief Visuospatial Memory Test—Revised (BVMT-R), the Wechsler Memory Scale—Third Edition: Spatial Span task (WMS-III-SS) forward and backwards, University of Maryland Letter-Number Span (LNS), the Brief Assessment of Cognition in Schizophrenia: Symbol Coding task (BACS-SC) (35), the Animal Category Fluency Task (Category Fluency), the Trail Making Test: Part A task (Trail A), the Continuous Performance Test—Identical Pairs (CPT-IP) (36), and the Neuropsychological Assessment Battery—Mazes task (NAB Mazes). In addition to the MCCB, Seidman’s Auditory Continuous Performance Test (A-CPT) was administered to assess auditory-verbal vigilance and sustained concentration under 3 conditions that included auditory-verbal vigilance alone (QA condition), auditory-verbal vigilance with a higher working memory demand (Q3A MEM), and auditory-verbal vigilance with both a higher working memory demand and the presence of interference (Q3A INT).

A few experimental tests were also included in the battery, including the Babble Test, which involved auditory perception of discrete words and/or phrases in the midst of a crowd of voices, and the University of Pennsylvania Smell Identification Test (UPSIT) (37), which involved olfactory identification of discrete odors in a multiple-choice format. A third experimental test involving paired-associate learning was only administered to portions of the sample and is not included for analysis here.

There was a baseline cognition visit, followed by a 12-month and a 24-month follow-up visit for most variables. Other variables were assessed more than three times.

### Statistical Analyses

Demographic comparisons between the groups (HC, CHR-C, CHR-R, CHR-NCNR), were calculated using a Kruskal-Wallis test for age and chi-squared tests for education (self and mother), sex, and medication status (i.e., currently taking anti-psychotic medication or not) at the time of the neuropsychological assessment.

Group differences in cognitive tests at baseline were evaluated using the Analysis of covariance (ANCOVA) with age, sex, income, and years of education (self and mother) as covariates. When ANCOVA assumptions were violated, rank transformations were used to address issues with normality, and robust ANCOVAs were used to address outcomes with severe outliers.

Paired t-tests were used to evaluate differences before and after conversion or remission on all of the cognitive tests. *“Before visits”* were defined as the *“Baseline visit,”* and *“After conversion,”* and were identified as the visit after conversion. *“Remission visits”* referred to the neurocognitive test battery data obtained at the next scheduled visit in participants who remitted. Linear models assessed the association between time since conversion and after conversion cognitive outcomes for converters to rule out a potential confounding effect of time since conversion and the cognitive assessment on associations between before and after conversion comparisons. The linear mixed-effects model (lmer) part of the lme4 package was fit to the data using a maximum likelihood estimation, which is a method of estimating the parameters of a statistical model by maximizing the likelihood function, and included age, sex, income, education (self and mother), and group*visit as fixed effects and a random intercept for subject IDs and slope for visit. The response variable was the cognitive test used for each of the models. Visit, or when the participants came in for an evaluation, was considered here as the time variable, which was either a baseline, 12-month, 24-month, or a corresponding converter visit. Two sets of sensitivity analyses were conducted, assessing differences in performance on cognitive tests at baseline and longitudinally, while accounting for the effects of anti-psychotic medications. As controls were excluded from the study if they were prescribed antipsychotic medication, they were not included in these analyses. Thus, CHR-NCNR were selected as the reference group for the lmer, and their cognitive trajectories were compared to other CHR groups. Analyses were conducted using RStudio version 4.2.0.

## Results

### Demographics

The groups (HC, CHR-C, CHR-R, and CHR-NCNR) differed significantly in age (p <.001), income (p < 0.01), education (self (p<0.001) and mother (p=0.02)). The HCs were older, had more years of education, and had higher income than the other groups. The groups did not differ by sex (Table 1).

**Table 1:** Demographics of CHR and Control participants in NAPLS-2 by converter status.

|  | HC<br>(N=265) | CHR-NCNR(N=384) | CHR-R (N=216) | CHR-C (N=89) | DF | *X*<sup>2</sup>/H | p | ηp<sup>2</sup>/V |
|---|---|---|---|---|---|---|---|---|
| Age+ | 19.775 (4.716) | 18.693 (4.371) | 18.169 (4.253) | 18.103 (3.628) | 3 | 19.6 | < 0.001 | 0.021 |
| Sex |  |  |  |  | 3 | 3.60 | 0.31 | 0.025 |
| Male | 137 (51.7%) | 221 (57.6%) | 122 (56.5%) | 55 (61.8%) |  |  |  |  |
| Female | 128 (48.3%) | 163 (42.4%) | 94 (43.5%) | 34 (38.2%) |  |  |  |  |
| Income |  |  |  |  | 18 | 38.6 | 0.003 | 0.085 |
| Less than \$10, 000 | 20 (7.5%) | 37 (9.7%) | 12 (5.6%) | 11 (12.5%) |  |  |  |  |
| \$10, 000 to \$19, 999 | 19 (7.2%) | 38 (10.0%) | 24 (11.1%) | 2 (2.3%) |  |  |  |  |
| \$20, 000 to \$39, 999 | 32 (12.1%) | 46 (12.1%) | 21 (9.7%) | 12 (13.6%) |  |  |  |  |
| \$40, 000 to \$59, 999 | 32 (12.1%) | 35 (9.2%) | 24 (11.1%) | 13 (14.8%) |  |  |  |  |
| \$60, 000 to \$99, 999 | 53 (20.0%) | 45 (11.8%) | 39 (18.1%) | 17 (19.3%) |  |  |  |  |
| \$100, 000 and above | 67 (25.3%) | 76 (19.9%) | 56 (25.9%) | 19 (21.6%) |  |  |  |  |
| Don't know or refused | 42 (15.8%) | 104 (27.3%) | 40 (18.5%) | 14 (15.9%) |  |  |  |  |
| Highest Level of Education |  |  |  |  | 15 | 101 | < 0.001 | 0.174 |
| Some high school | 93 (35.1%) | 203 (53.1%) | 121 (56.0%) | 48 (53.9%) |  |  |  |  |
| High school | 89 (33.6%) | 137 (35.9%) | 78 (36.1%) | 37 (41.6%) |  |  |  |  |
| College | 24 (9.1%) | 15 (3.9%) | 5 (2.3%) | 2 (2.2%) |  |  |  |  |
| Technical school | 3 (1.1%) | 11 (2.9%) | 0 (0.0%) | 0 (0.0%) |  |  |  |  |
| University | 46 (17.4%) | 13 (3.4%) | 11 (5.1%) | 2 (2.2%) |  |  |  |  |
| Graduate school | 10 (3.8%) | 3 (0.8%) | 1 (0.5%) | 0 (0.0%) |  |  |  |  |
| Highest Level of Mother's Education |  |  |  |  | 21 | 36.4 | 0.02 | 0.074 |
| Some primary school | 1 (0.4%) | 7 (1.9%) | 5 (2.3%) | 2 (2.3%) |  |  |  |  |
| Completed primary school | 3 (1.1%) | 9 (2.4%) | 3 (1.4%) | 1 (1.1%) |  |  |  |  |
| Some high school | 7 (2.7%) | 30 (8.0%) | 16 (7.5%) | 9 (10.2%) |  |  |  |  |
| Completed high school | 35 (13.3%) | 67 (17.9%) | 40 (18.8%) | 9 (10.2%) |  |  |  |  |
| Some college/technical school/undergraduate | 55 (20.8%) | 85 (22.7%) | 35 (16.4%) | 22 (25.0%) |  |  |  |  |
| Complete college/technical school/undergraduate | 96 (36.4%) | 118 (31.5%) | 75 (35.2%) | 25 (28.4%) |  |  |  |  |
| Some graduate/professional school | 7 (2.7%) | 12 (3.2%) | 8 (3.8%) | 4 (4.5%) |  |  |  |  |
| Completed graduate/professional school | 60 (22.7%) | 47 (12.5%) | 31 (14.6%) | 16 (18.2%) |  |  |  |  |

*Note: +: Kruskal-Wallis Test, HC: Healthy Controls, CHR-NCNR: Clinical High Risk Non-Converter Non-Remitter, CHR-R: Clinical High Risk - Remitter, CHR-C: Clinical High Risk - Converter, DF: Degrees of Freedom, X<sup>2</sup>/H: Pearson’s X<sup>2</sup> test statistic for categorical variables / Kruskal-Wallis H statistic for continuous variables, p: p-value, ηp<sup>2</sup>: Partial Eta Squared; V: Cramers-V.*

### Neurocognition

#### Baseline comparisons

HCs performed better on all cognitive measures versus all CHR groups, with the exception of the WRAT-4, WASI-BD, NAB Mazes, UPSIT, and BSIT, which did not differ between any of the groups. CHR-NCNR performed worse than the CHR-R on the Babble Total test (p=0.029). CHR-C performed significantly worse than CHR-NCNR in a variety of measures, including Category Fluency, BVMT-R, HVLT-R, and Trails A. CHR-C also performed significantly worse than CHR-R in the Babble test (longest word) (Table 2, Figure 1a-c). Sensitivity analyses suggest that use of antipsychotics do not account for variability in cognitive outcomes at baseline (Table ST1).

**Figure 1a-c:**
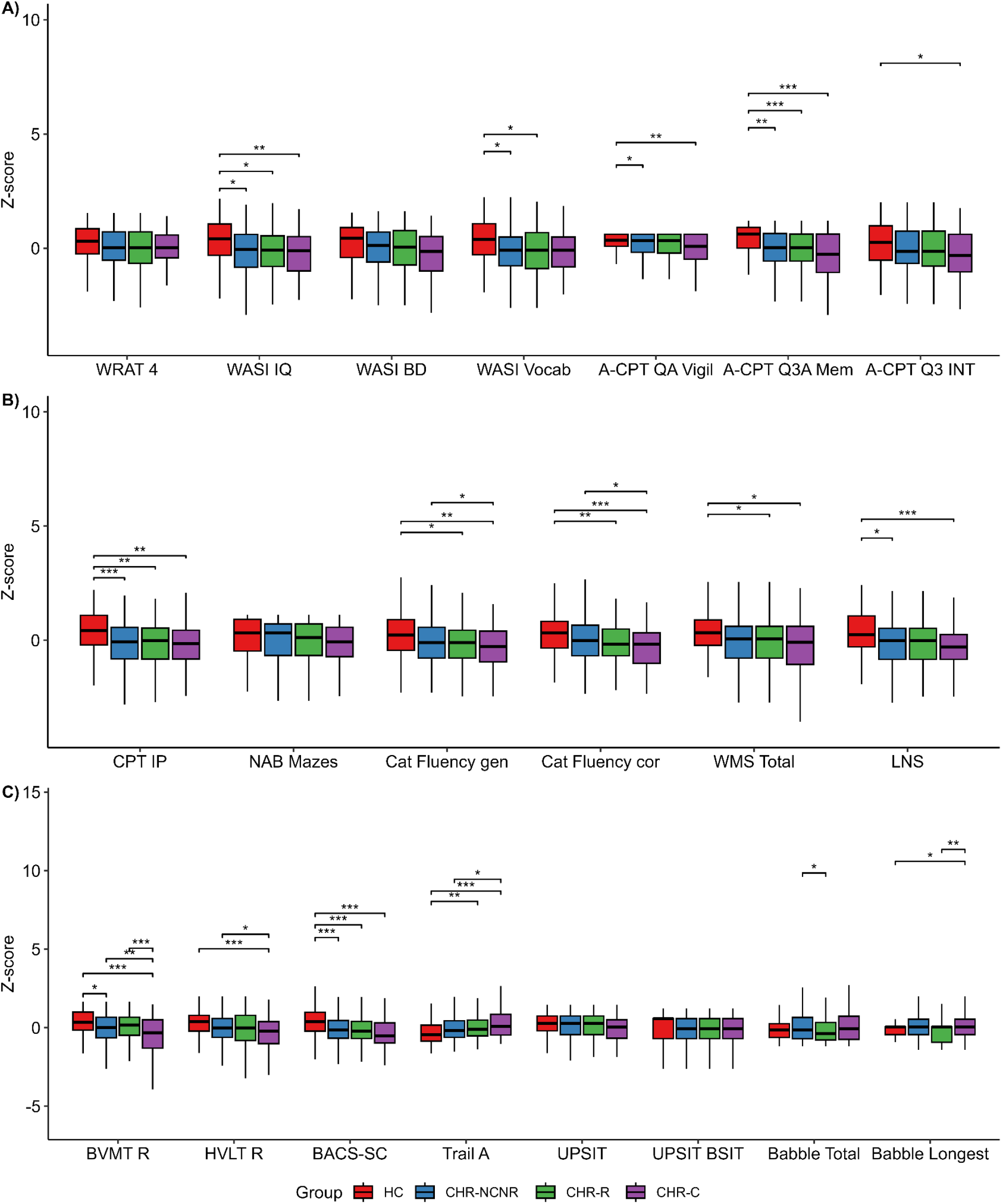
Baseline differences of cognitive assessment performance across converter status groups. Note: ***: p <0.001, **: p < 0.01, *: p <0.05. HC: Healthy Controls, CHR-NCNR: Clinical High Risk Non-Converter Non-Remitter, CHR-R: Clinical High Risk - Remitter; CHR-C: Clinical High Risk - Converter; WRAT-4: Wide Range Achievement Test, 4th Edition – Reading, WASI IQ: WASI-II Full Scale IQ (2-subtest version), WASI BD: WASI-II Block Design, WASI Vocab: WASI-II Vocabulary, A-CPT QA Vigil: Auditory Continuous Performance Test, QA Vigilance, A-CPT Q3A Mem: Auditory Continuous Performance Test, Q3A Memory, A-CPT Q3 INT: Auditory Continuous Performance Test, Q3 Interference, CPT-IP: Continuous Performance Test – Identical Pairs, NAB Mazes: Neuropsychological Assessment Battery – Mazes, Category Fluency gen/cor: Category Fluency – number of words generated / correct words, WMS Total: Wechsler Memory Scale – Total Score, LNS: University of Maryland Letter-Number Sequencing, BVMT R: Brief Visuospatial Memory Test - Revised, HVLT R: Hopkins Verbal Learning Test - Revised, BACS-SC: Brief Assessment of Cognition in Schizophrenia – Symbol Coding, Trail A: Trail Making Test: Part A, UPSIT/UPSIT BSIT: University of Pennsylvania Smell Identification Test, Babble Total: Total Word Count in Babble Task, Babble Longest: Longest Continuous Phrase in Babble Task.

**Table 2:** Baseline differences in cognitive assessments across converter status groups.

| Outcome | HC (■) | CHR-NCNR (○) | CHR-R (⊗) | CHR-C (●) | *F/X*<sup>2</sup> | *df* | *ηp²/V* | *p* |
|---|---|---|---|---|---|---|---|---|
| WRAT-4# | 60.39 (6.36) | 58.27 (7.41) | 57.79 (8.29) | 58.78 (5.57) | 0.04 | 3.00 | 0.03 | ■○⊗● = 0.99 |
| WASI-IQ | 111 (14.14) | 103.72 (15.33) | 104.25 (15.52) | 102.06 (14.59) | 4.78 | 3.00 | 0.06 | ■○⊗● = **<br>■○ = *<br>■⊗ = *<br>■● = ** |
| WASI-BD | 49.1 (14.42) | 45.74 (15.35) | 44.9 (15.53) | 42.61 (15.62) | 1.31 | 3.00 | 0.02 | ■○⊗● = 0.26 |
| WASI-Vocab | 59.82 (9.48) | 54.51 (10.39) | 54.37 (10.57) | 54.18 (8.84) | 3.59 | 3.00 | 0.08 | ■○⊗● = *<br>■○ = *<br>■⊗ = * |
| A-CPT QA Vigil#+ | 97.24 (4.93) | 95.22 (7.1) | 94.41 (9.3) | 92.32 (10.35) | 18.86 | 3.00 | 0.08 | ■○⊗● = ***<br>■○ = *<br>■● = ** |
| A-CPT Q3A Mem<br># | 87.58 (10.82) | 82.12 (14.03) | 81.31 (14.06) | 76.01 (18.89) | 10.33 | 3.00 | 0.06 | ■○⊗● = ***<br>■○ = **<br>■⊗ = ***<br>■● = *** |
| A-CPT Q3 INT | 61.03 (20.71) | 56.1 (20.47) | 56.37 (21.78) | 51.04 (23.26) | 2.86 | 3.00 | 0.02 | ■○⊗● = *<br>■● = * |
| CPT IP | 2.82 (0.73) | 2.4 (0.75) | 2.4 (0.79) | 2.34 (0.84) | 7.56 | 3.00 | 0.08 | ■○⊗● = ***<br>■○ = ***<br>■⊗ = **<br>■● = ** |
| NAB Mazes# | 20.95 (4.8) | 20.33 (5.18) | 20.03 (5.13) | 19.69 (4.94) | 1.22 | 3.00 | 0.01 | ■○⊗● = 0.3 |
| Category Fluency gen | 26.11 (5.78) | 24.56 (6.14) | 23.7 (5.5) | 22.63 (5.65) | 6.12 | 3.00 | 0.04 | ■○⊗● = ***<br>■⊗ = *<br>■● = **<br>○● = * |
| Category Fluency cor | 25.66 (5.76) | 23.97 (5.99) | 23.01 (5.88) | 22.03 (5.77) | 7.10 | 3.00 | 0.04 | ■○⊗● = ***<br>■⊗ = **<br>■● = ***<br>○● = * |
| WMS Total | 17.73 (3.34) | 16.55 (3.66) | 16.41 (3.23) | 16.06 (4.38) | 3.75 | 3.00 | 0.03 | ■○⊗● = *<br>■⊗ = *<br>■● = * |
| LNS | 16.31 (3.37) | 14.72 (3.52) | 14.76 (3.79) | 13.65 (4.07) | 5.97 | 3.00 | 0.06 | ■○⊗● = ***<br>■○ = *<br>■● = *** |
| BVMT-R | 27.55 (5.4) | 25.57 (6.05) | 25.83 (6.06) | 22.97 (6.97) | 9.10 | 3.00 | 0.05 | ■○⊗● = ***<br>■○ = *<br>■● = ***<br>○● = **<br>⊗● = *** |
| HVLT-R | 27.44 (4.22) | 25.77 (4.96) | 25.61 (5.45) | 24.32 (5.27) | 5.58 | 3.00 | 0.04 | ■○⊗● = ***<br>■● = ***<br>○● = * |
| BACS Symbol Coding# | 64.27 (12.91) | 57.45 (12.67) | 57.16 (13.56) | 53.48 (12.08) | 13.21 | 3.00 | 0.09 | ■○⊗● = ***<br>■○ = ***<br>■⊗ = ***<br>■● = *** |
| Trails A# | 26.47 (10.59) | 29.53 (11.91) | 30.22 (11.97) | 32.35 (11.17) | 7.15 | 3.00 | 0.05 | ■○⊗● = ***<br>■⊗ = **<br>■● = ***<br>○● = * |
| UPSIT#+ | 34.56 (3.53) | 33.7 (4.35) | 33.54 (4.52) | 33.01 (4.8) | 3.01 | 3.01 | 0.03 | ■○⊗● = 0.39 |
| BSIT# | 10.3 (1.44) | 9.99 (1.56) | 10.02 (1.71) | 10 (1.57) | 1.12 | 0.73 | 0.01 | ■○⊗● = 0.53 |
| Babble total words#+ | 13.94 (9.96) | 16 (14.36) | 13.07 (11.76) | 16.6 (12.86) | 8.97 | 8.94 | 0.06 | ■○⊗● = *<br>○⊗= * |
| Babble Longest#+ | 2.91 (2.3) | 2.9 (1.82) | 2.7 (2.09) | 3.31 (1.98) | 11.25 | 11.25 | 0.06 | ■○⊗● = *<br>■● = *<br>⊗● = ** |

*Note #: Rank order ANCOVA+: Robust ANCOVA: +: Kruskal-Wallis Test, HC: Healthy Controls, CHR-NCNR: Clinical High Risk Non-Converter Non-Remitter, CHR-R: Clinical High Risk - Remitter; CHR-C: Clinical High Risk - Converter; ηp²: Partial Eta Squared; V: Cramers-V. ■: HC; ○: CHR-NCNR; ⊗: CHR-R; ●: CHR-C. \*\*\*: p <0.001, \*\*: p < 0.01, \*: p <0.05*. WRAT-4: *Wide Range Achievement Test, 4th Edition – Reading*, WASI IQ: *WASI-II Full Scale IQ (2-subtest version)*, WASI BD: *WASI-II Block Design,* WASI Vocab: *WASI-II Vocabulary*, A-CPT QA Vigil: *Auditory Continuous Performance Test, QA Vigilance*, A-CPT Q3A Mem: *Auditory Continuous Performance Test, Q3A Memory*, A-CPT Q3 INT: *Auditory Continuous Performance Test, Q3 Interference*, CPT-IP: *Continuous Performance Test – Identical Pairs*, NAB Mazes: *Neuropsychological Assessment Battery – Mazes*, Category Fluency gen/cor: *Category Fluency – number of words generated / correct words,* WMS Total: *Wechsler Memory Scale – Total Score*, LNS: *University of Maryland Letter-Number Sequencing*, BVMT R: *Brief Visuospatial Memory Test - Revised,* HVLT R: *Hopkins Verbal Learning Test - Revised*, BACS-SC: *Brief Assessment of Cognition in Schizophrenia – Symbol Coding*, Trail A: *Trail Making Test: Part A*, UPSIT/UPSIT BSIT: *University of Pennsylvania Smell Identification Test*, Babble Total: *Total Word Count in Babble Task,* Babble Longest*: Longest Continuous Phrase in Babble Task*.

#### Comparisons before and after conversion

Four of 21 measures showed performance improvements in the CHR-C group after conversion to psychosis, including WASI Block Design (q< 0.001), A-CPT Q3A Memory (q=0.026), A-CPT Q3A Interference (q=0.026), and Trails A (q< 0.001) (Table 3, Figure 2). The average time between conversion and the post-conversion neuropsychological assessment was 123.61 days (SD = 118.01).

**Figure 2:**
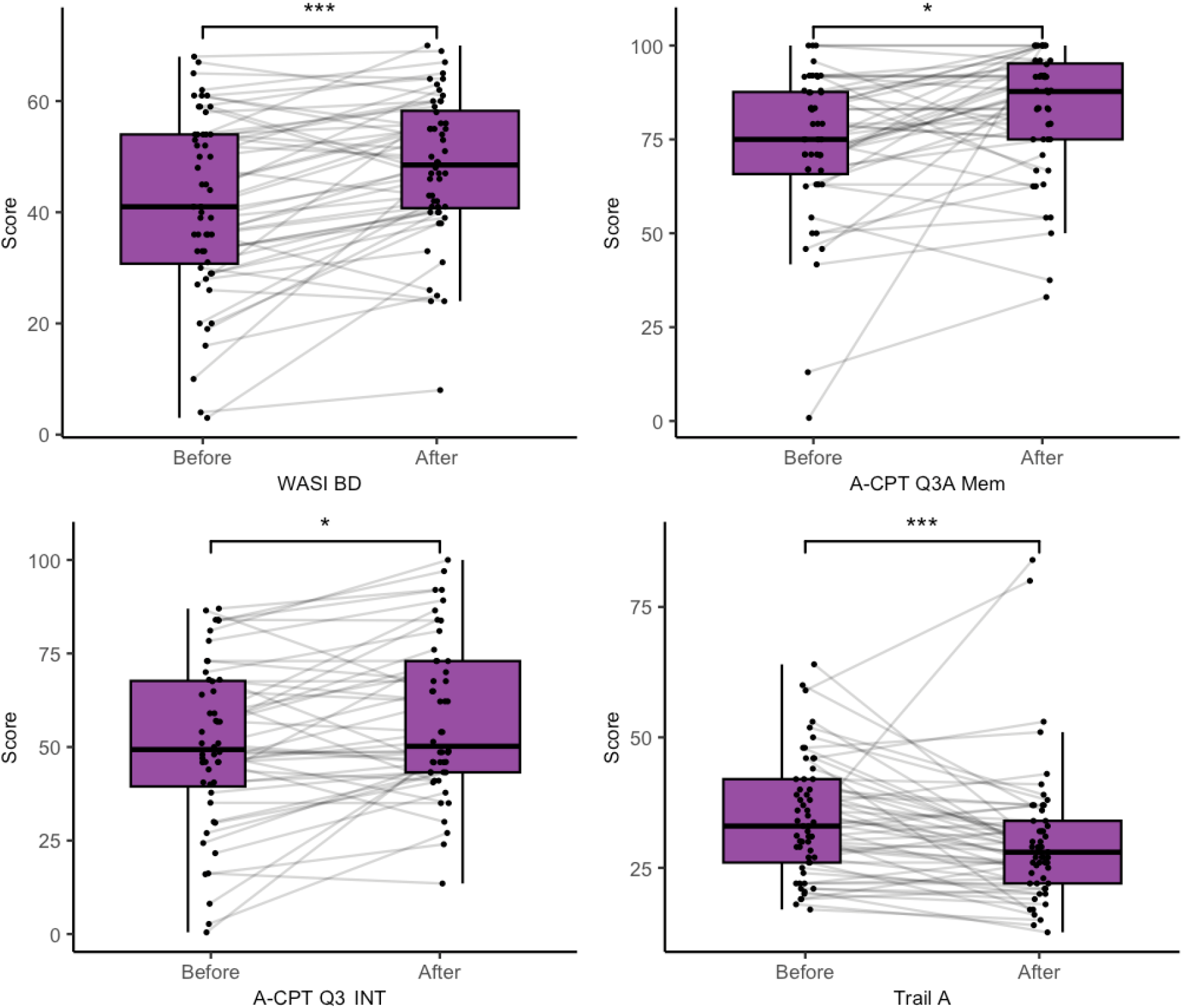
Significant differences in cognitive assessment performance before and after conversion visits. WASI BD: WASI-II Block Design, A-CPT Q3A Mem: Auditory Continuous Performance Test, Q3A Memory, A-CPT Q3 INT: Auditory Continuous Performance Test, Q3 Interference, Trail A: Trail Making Test Part A

**Table 3:**
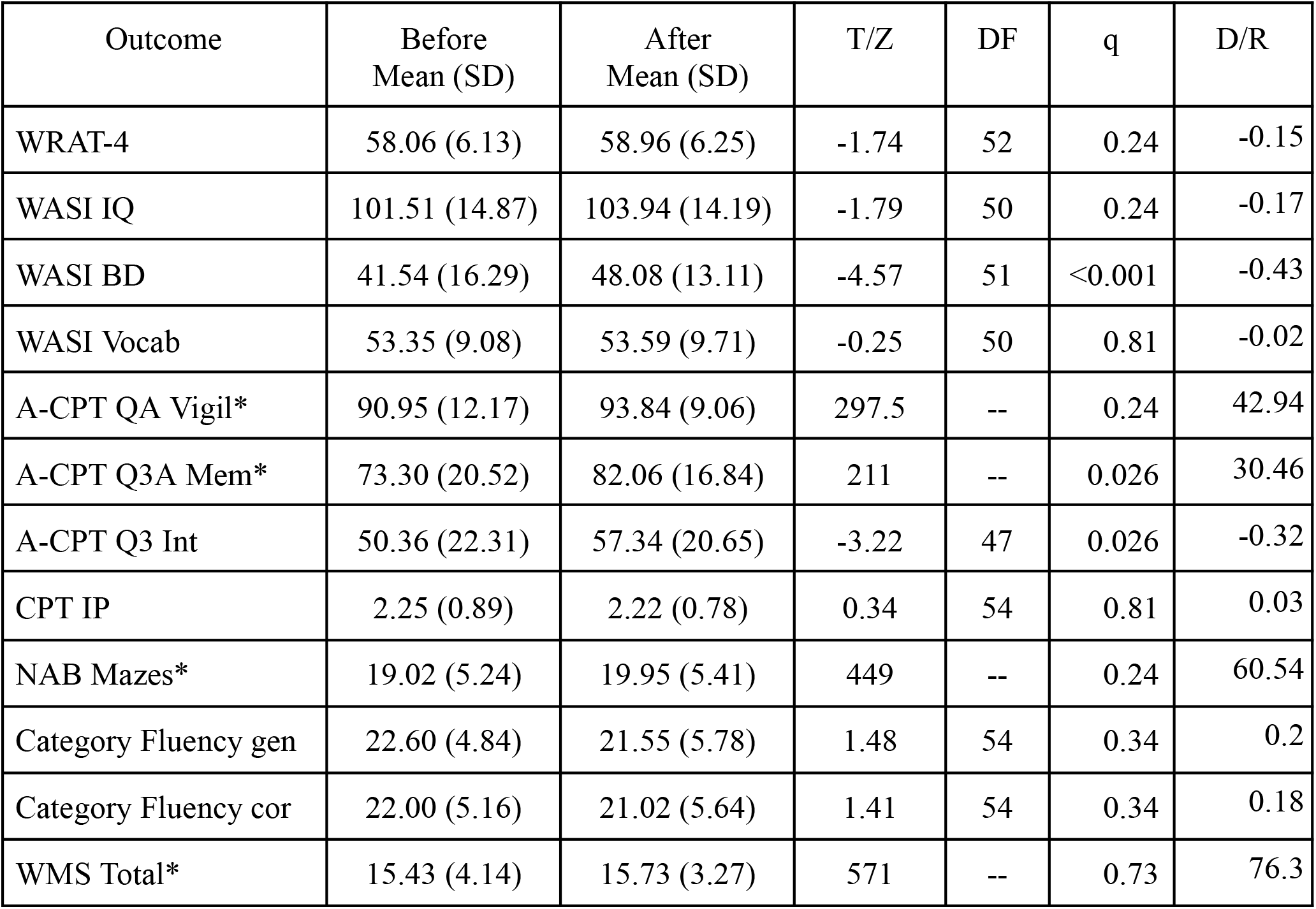
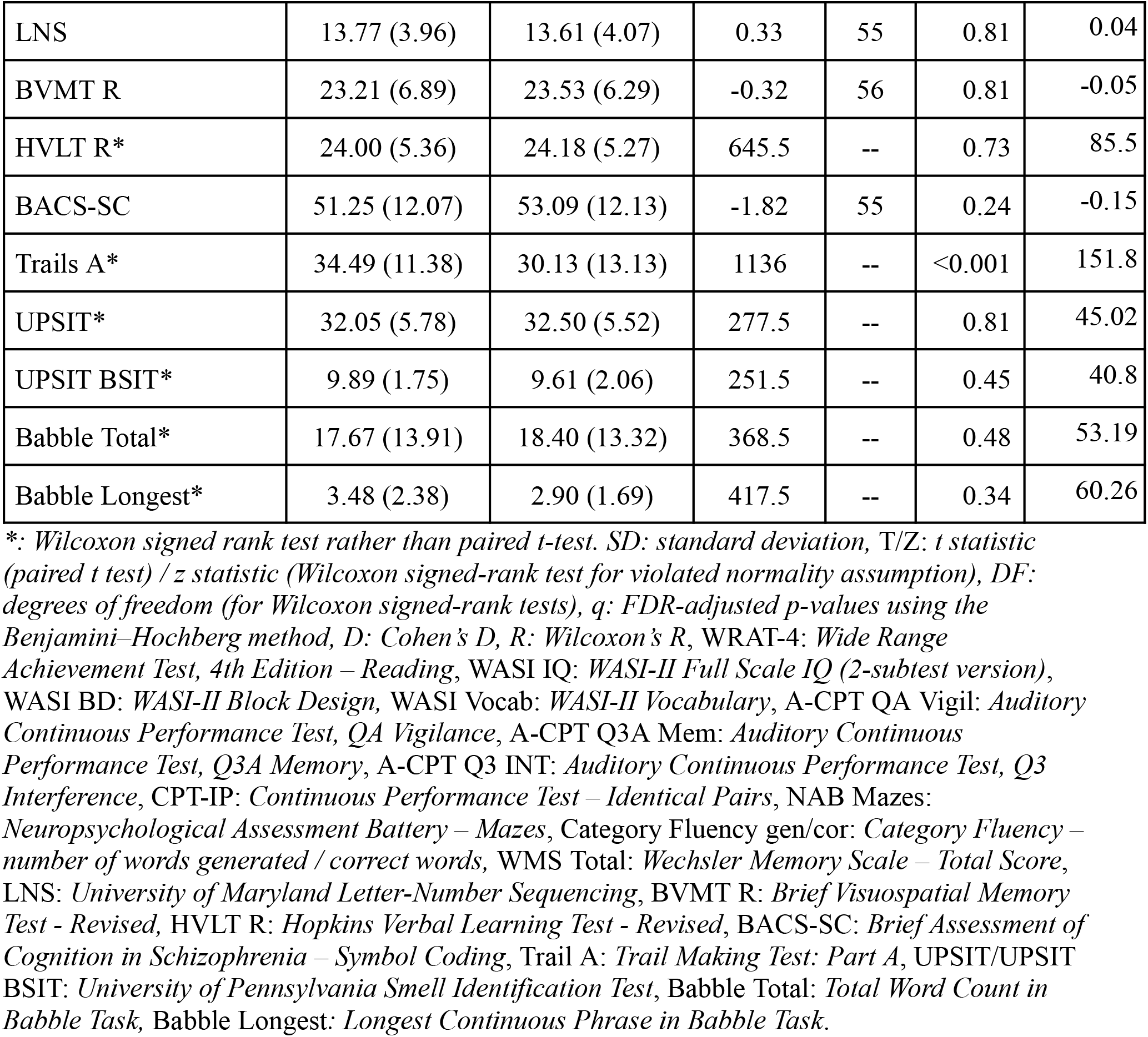
Paired T-Tests comparisons of performance on cognitive assessments before and after conversion visits.

| Outcome | Before<br>Mean (SD) | After<br>Mean (SD) | T/Z | DF | q | D/R |
|---|---|---|---|---|---|---|
| WRAT-4 | 58.06 (6.13) | 58.96 (6.25) | -1.74 | 52 | 0.24 | -0.15 |
| WASI IQ | 101.51 (14.87) | 103.94 (14.19) | -1.79 | 50 | 0.24 | -0.17 |
| WASI BD | 41.54 (16.29) | 48.08 (13.11) | -4.57 | 51 | <0.001 | -0.43 |
| WASI Vocab | 53.35 (9.08) | 53.59 (9.71) | -0.25 | 50 | 0.81 | -0.02 |
| A-CPT QA Vigil* | 90.95 (12.17) | 93.84 (9.06) | 297.5 | -- | 0.24 | 42.94 |
| A-CPT Q3A Mem* | 73.30 (20.52) | 82.06 (16.84) | 211 | -- | 0.026 | 30.46 |
| A-CPT Q3 Int | 50.36 (22.31) | 57.34 (20.65) | -3.22 | 47 | 0.026 | -0.32 |
| CPT IP | 2.25 (0.89) | 2.22 (0.78) | 0.34 | 54 | 0.81 | 0.03 |
| NAB Mazes* | 19.02 (5.24) | 19.95 (5.41) | 449 | -- | 0.24 | 60.54 |
| Category Fluency gen | 22.60 (4.84) | 21.55 (5.78) | 1.48 | 54 | 0.34 | 0.2 |
| Category Fluency cor | 22.00 (5.16) | 21.02 (5.64) | 1.41 | 54 | 0.34 | 0.18 |
| WMS Total* | 15.43 (4.14) | 15.73 (3.27) | 571 | -- | 0.73 | 76.3 |
| LNS | 13.77 (3.96) | 13.61 (4.07) | 0.33 | 55 | 0.81 | 0.04 |
| BVMT R | 23.21 (6.89) | 23.53 (6.29) | -0.32 | 56 | 0.81 | -0.05 |
| HVLT R* | 24.00 (5.36) | 24.18 (5.27) | 645.5 | -- | 0.73 | 85.5 |
| BACS-SC | 51.25 (12.07) | 53.09 (12.13) | -1.82 | 55 | 0.24 | -0.15 |
| Trails A* | 34.49 (11.38) | 30.13 (13.13) | 1136 | -- | <0.001 | 151.8 |
| UPSIT* | 32.05 (5.78) | 32.50 (5.52) | 277.5 | -- | 0.81 | 45.02 |
| UPSIT BSIT* | 9.89 (1.75) | 9.61 (2.06) | 251.5 | -- | 0.45 | 40.8 |
| Babble Total* | 17.67 (13.91) | 18.40 (13.32) | 368.5 | -- | 0.48 | 53.19 |
| Babble Longest* | 3.48 (2.38) | 2.90 (1.69) | 417.5 | -- | 0.34 | 60.26 |

*\*: Wilcoxon signed rank test rather than paired t-test. SD: standard deviation,* T/Z: *t statistic (paired t test) / z statistic (Wilcoxon signed-rank test for violated normality assumption), DF: degrees of freedom (for Wilcoxon signed-rank tests), q: FDR-adjusted p-values using the Benjamini–Hochberg method, D: Cohen's D, R: Wilcoxon's R*, WRAT-4: *Wide Range Achievement Test, 4th Edition – Reading*, WASI IQ: *WASI-II Full Scale IQ (2-subtest version)*, WASI BD: *WASI-II Block Design,* WASI Vocab: *WASI-II Vocabulary*, A-CPT QA Vigil: *Auditory Continuous Performance Test, QA Vigilance*, A-CPT Q3A Mem: *Auditory Continuous Performance Test, Q3A Memory*, A-CPT Q3 INT: *Auditory Continuous Performance Test, Q3 Interference*, CPT-IP: *Continuous Performance Test – Identical Pairs*, NAB Mazes: *Neuropsychological Assessment Battery – Mazes*, Category Fluency gen/cor: *Category Fluency – number of words generated / correct words,* WMS Total: *Wechsler Memory Scale – Total Score*, LNS: *University of Maryland Letter-Number Sequencing*, BVMT R: *Brief Visuospatial Memory Test - Revised,* HVLT R: *Hopkins Verbal Learning Test - Revised*, BACS-SC: *Brief Assessment of Cognition in Schizophrenia – Symbol Coding*, Trail A: *Trail Making Test: Part A*, UPSIT/UPSIT BSIT: *University of Pennsylvania Smell Identification Test*, Babble Total: *Total Word Count in Babble Task,* Babble Longest*: Longest Continuous Phrase in Babble Task.*

#### Comparisons before and after remission

The CHR-R group showed significant improvement in 14 of 21 measures, including WRAT-4 (q<0.001), WASI IQ (q<0.001), WASI Block Design (q < 0.001), WASI Vocabulary (q<0.001), A-CPT QA Vigilance (q <0.01), A-CPT Q3A Memory (q <0.01), A-CPT Q3A Interference (q < 0.001), CPT IP (q < 0.001), NAB Mazes (q <0.001), Wechsler Memory Scale III, Spatial Span (q <0.001), Letter-Number Sequencing (q <0.001), BACS Symbol Coding (q <0.001), Trails A (q <0.001), and Babble Total (q <0.001) from before to after remission. No change was observed in the other neuropsychological assessments (Table 4, Figure 3).

**Figure 3:**
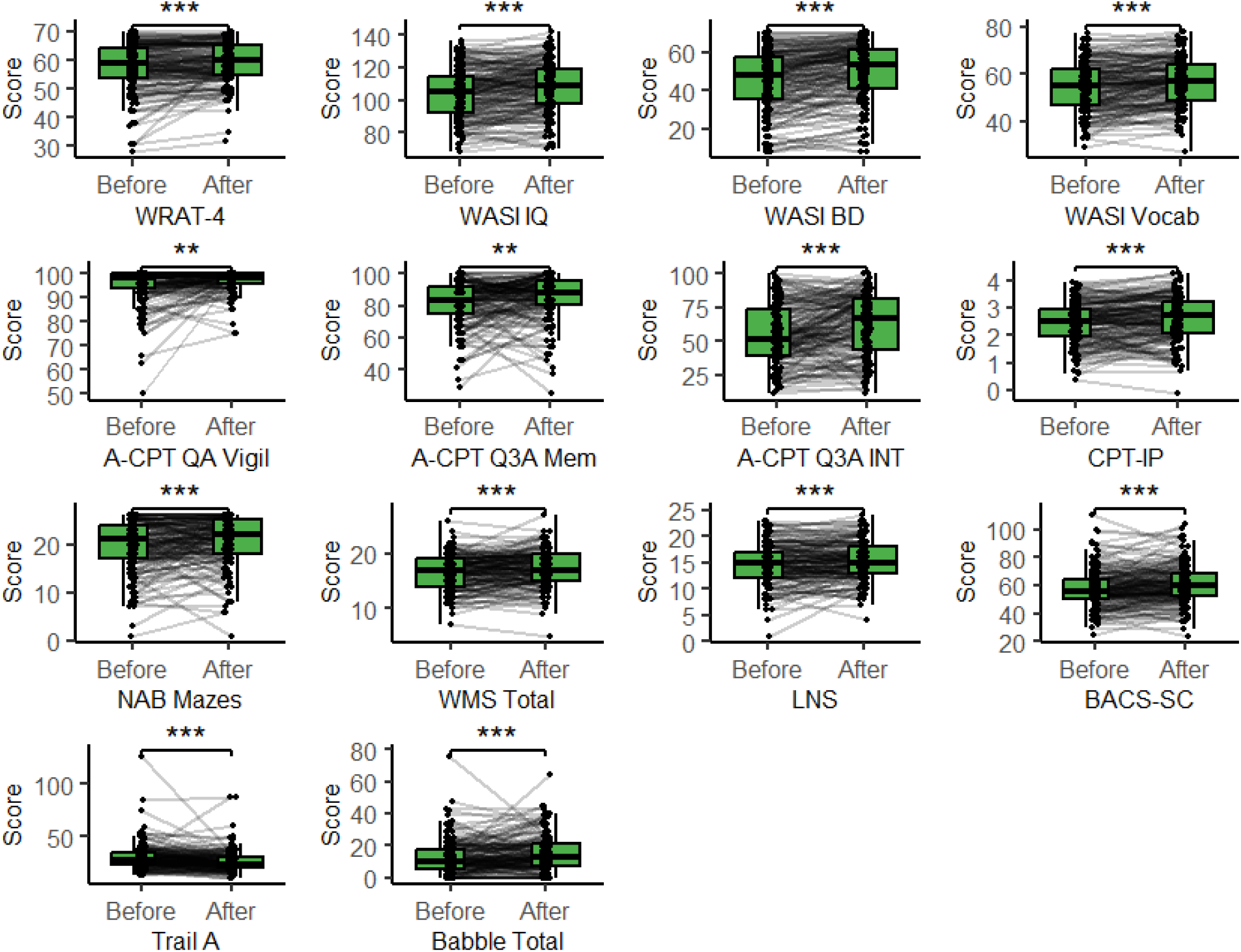
Significant differences in cognitive assessment performance before and after remission visits WRAT-4: Wide Range Achievement Test, 4th Edition – Reading, WASI IQ: WASI-II Full Scale IQ (2-subtest version), WASI BD: WASI-II Block Design, WASI Vocab: WASI-II Vocabulary, A-CPT QA Vigil: Auditory Continuous Performance Test, QA Vigilance, A-CPT Q3A Mem: Auditory Continuous Performance Test, Q3A Memory, A-CPT Q3 INT: Auditory Continuous Performance Test, Q3 Interference, CPT-IP: Continuous Performance Test – Identical Pairs, NAB Mazes: Neuropsychological Assessment Battery – Mazes, WMS Total: Wechsler Memory Scale – Total Score, LNS: University of Maryland Letter-Number Sequencing, BACS-SC: Brief Assessment of Cognition in Schizophrenia – Symbol Coding, Trail A: Trail Making Test Part A, Babble Total: Total Word Count in Babble Task.

**Table 4:** Paired T-Tests comparisons of performance on cognitive assessments before and after remission visits.

| Outcome | Before | After | T/Z | DF | q | D/R |
|---|---|---|---|---|---|---|
| WRAT-4* | 57.67 (8.44) | 59.27 (6.99) | 2098.5 | -- | <0.001 | 164.87 |
| WASI IQ | 104.36 (15.57) | 107.99 (15.39) | -5.89 | 161 | <0.001 | -0.23 |
| WASI BD* | 44.94 (15.71) | 50.15 (15.18) | 1741.5 | -- | <0.001 | 136.83 |
| WASI Vocab | 54.42 (10.30) | 57.07 (9.99) | -5.76 | 161 | <0.001 | -0.26 |
| A-CPT QA Vigil* | 95.14 (7.52) | 97.24 (4.37) | 2078.5 | -- | <0.01 | 165.36 |
| A-CPT Q3A Mem* | 81.77 (13.81) | 85.19 (13.89) | 3504.5 | -- | <0.01 | 278.8 |
| A-CPT Q3 INT* | 55.22 (21.66) | 62.39 (22.61) | 3033 | -- | <0.001 | 242.83 |
| CPT IP* | 2.45 (0.77) | 2.63 (0.82) | 3855 | -- | <0.001 | 304.76 |
| NAB Mazes* | 19.96 (5.22) | 21.03 (4.90) | 3017.5 | -- | <0.001 | 234.2 |
| Category Fluency gen | 23.58 (5.42) | 23.81 (5.33) | -0.6 | 166 | 0.71 | -0.04 |
| Category Fluency cor | 23.19 (5.37) | 23.20 (5.69) | -0.04 | 166 | 0.97 | 0 |
| WMS Total* | 16.51 (3.14) | 17.30 (3.27) | 3785 | -- | <0.001 | 293.77 |
| LNS | 14.72 (3.93) | 15.42 (3.60) | -3.51 | 164 | <0.001 | -0.19 |
| BVMT R* | 25.73 (6.23) | 26.37 (5.62) | 5180 | -- | 0.38 | 403.26 |
| HVLT R* | 25.79 (5.26) | 26.12 (4.73) | 5069 | -- | 0.38 | 391.08 |
| BACS-SC* | 57.51 (14.01) | 60.93 (14.04) | 3841 | -- | <0.001 | 297.23 |
| Trail A* | 30.52 (12.78) | 26.42 (10.47) | 9663 | -- | <0.001 | 749.99 |
| UPSIT* | 33.62 (4.03) | 33.90 (4.03) | 2692 | -- | 0.38 | 234.31 |
| UPSIT BSIT* | 9.99 (1.67) | 9.96 (1.68) | 2633 | -- | 0.79 | 230.05 |
| Babble Total* | 12.46 (11.02) | 15.36 (11.52) | 3305.5 | -- | <0.001 | 262.97 |
| Babble Longest* | 2.69 (2.22) | 2.74 (2.09) | 2746 | -- | 0.66 | 218.46 |

\*: *Wilcoxon signed rank test rather than paired t-test.* T/Z = *T statistic (paired t test) / Z statistic (Wilcoxon signed-rank test for violated normality assumption); DF = Degrees of Freedom (for Wilcoxon signed-rank tests); q = FDR-adjusted p-values using the Benjamini–Hochberg method, D: Cohen’s D, R: Wilcoxon’s R;* WRAT-4: *Wide Range Achievement Test, 4th Edition – Reading*, WASI IQ: *WASI-II Full Scale IQ (2-subtest version)*, WASI BD: *WASI-II Block Design,* WASI Vocab: *WASI-II Vocabulary*, A-CPT QA Vigil: *Auditory Continuous Performance Test, QA Vigilance*, A-CPT Q3A Mem: *Auditory Continuous Performance Test, Q3A Memory*, A-CPT Q3 INT: *Auditory Continuous Performance Test, Q3 Interference*, CPT-IP: *Continuous Performance Test – Identical Pairs*, NAB Mazes: *Neuropsychological Assessment Battery – Mazes*, Category Fluency gen/cor: *Category Fluency – number of words generated / correct words,* WMS Total: *Wechsler Memory Scale – Total Score*, LNS: *University of Maryland Letter-Number Sequencing*, BVMT R: *Brief Visuospatial Memory Test - Revised,* HVLT R: *Hopkins Verbal Learning Test - Revised*, BACS-SC: *Brief Assessment of Cognition in Schizophrenia – Symbol Coding*, Trail A: *Trail Making Test: Part A*, UPSIT/UPSIT BSIT: *University of Pennsylvania Smell Identification Test*, Babble Total: *Total Word Count in Babble Task,* Babble Longest*: Longest Continuous Phrase in Babble Task*.

No associations were observed between these cognitive outcomes and the interval between the conversion episode and the post-conversion neuropsychological assessment (Table ST2).

### Longitudinal comparisons

#### Controls vs CHR-NCNR, Converters, Remitters

Comparisons showed that CHR-NCNR and CHR-R performance improved significantly more than HC on the A-CPT QA vigilance test, while CHR-C remained lower than HC over time. CHR-C did show a significant improvement compared to HC on the A-CPT Q3A memory test. No other significant Group X Time interactions between the HC and the other groups were observed on any of the tests (Table 5, Figure 4). Sensitivity analyses suggested that use of antipsychotics also do not account for variability in cognitive outcomes (Table ST3). No baseline difference in cognition were observed between those who dropped out and those who were followed up except in CPT IP (Table ST4).

**Figure 4:**
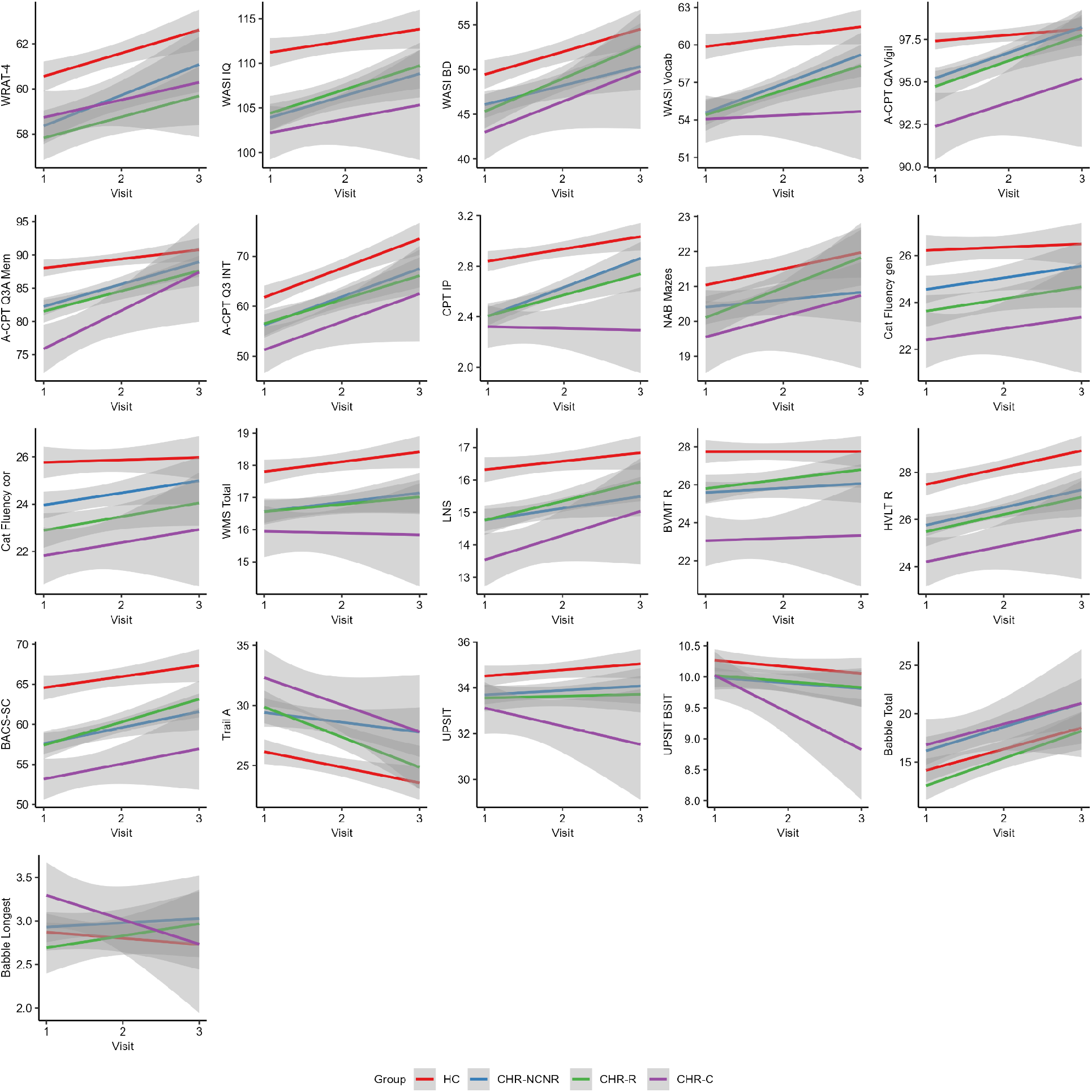
Longitudinal Cognitive Outcomes between Healthy Controls and converter status groups. HC: Healthy Controls, CHR-NCNR: Clinical High Risk Non-Converter Non-Remitter, CHR-R: Clinical High Risk - Remitter; CHR-C: Clinical High Risk - Converter; WRAT-4: Wide Range Achievement Test, 4th Edition – Reading, WASI IQ: WASI-II Full Scale IQ (2-subtest version), WASI BD: WASI-II Block Design, WASI Vocab: WASI-II Vocabulary, A-CPT QA Vigil: Auditory Continuous Performance Test, QA Vigilance, A-CPT Q3A Mem: Auditory Continuous Performance Test, Q3A Memory, A-CPT Q3 INT: Auditory Continuous Performance Test, Q3 Interference, CPT-IP: Continuous Performance Test – Identical Pairs, NAB Mazes: Neuropsychological Assessment Battery – Mazes, Category Fluency gen/cor: Category Fluency – number of words generated / correct words, WMS Total: Wechsler Memory Scale – Total Score, LNS: University of Maryland Letter-Number Sequencing, BVMT R: Brief Visuospatial Memory Test - Revised, HVLT R: Hopkins Verbal Learning Test - Revised, BACS-SC: Brief Assessment of Cognition in Schizophrenia – Symbol Coding, Trail A: Trail Making Test: Part A, UPSIT/UPSIT BSIT: University of Pennsylvania Smell Identification Test, Babble Total: Total Word Count in Babble Task, Babble Longest: Longest Continuous Phrase in Babble Task. Visit 1: Baseline, Visit 2: 12-month or converter visit after baseline, Visit 3: 24-month or converter visit after Visit 2.

**Table 5:** Linear mixed effect models assessing differences in longitudinal cognitive performance between Controls and Converter status groups.

| Outcome | Predictor | Estimate | SE | df | T | q |
|---|---|---|---|---|---|---|
| WRAT-4 | Visit | -1.01 | 0.37 | 1152.03 | -2.74 | 0.07 |
|  | CHR-NCNR*Visit | 0.03 | 0.23 | 985.33 | 0.15 | 0.95 |
|  | CHR-R*Visit | -0.03 | 0.22 | 966.47 | -0.14 | 0.95 |
|  | CHR-C*Visit | 0.1 | 0.41 | 965.91 | 0.24 | 0.95 |
| WASI IQ | Visit | -0.8 | 0.84 | 1086.23 | -0.95 | 0.5 |
|  | CHR-NCNR*Visit | 0.72 | 0.52 | 465.58 | 1.39 | 0.29 |
|  | CHR-R*Visit | 0.88 | 0.52 | 431.03 | 1.71 | 0.21 |
|  | CHR-C*Visit | 0.66 | 0.95 | 790.15 | 0.69 | 0.62 |
| WASI BD | Visit | 1.27 | 0.78 | 1050.99 | 1.63 | 0.23 |
|  | CHR-NCNR*Visit | -0.24 | 0.49 | 466.38 | -0.48 | 0.71 |
|  | CHR-R*Visit | 0.6 | 0.49 | 428.46 | 1.23 | 0.35 |
|  | CHR-C*Visit | 2.03 | 0.88 | 799.99 | 2.32 | 0.07 |
| WASI Vocab | Visit | -3.31 | 0.59 | 1098.64 | -5.59 | < 0.001 |
|  | CHR-NCNR*Visit | 0.44 | 0.39 | 480.52 | 1.13 | 0.42 |
|  | CHR-R*Visit | 0.8 | 0.39 | 446.99 | 2.08 | 0.12 |
|  | CHR-C*Visit | -0.01 | 0.69 | 790.04 | -0.02 | 0.98 |
| A-CPT QA Vigil | Visit | -0.29 | 0.51 | 1566.03 | -0.56 | 0.86 |
|  | CHR-NCNR*Visit | 1.02 | 0.4 | 1020.21 | 2.53 | < 0.05 |
|  | CHR-R*Visit | 1.1 | 0.42 | 856.84 | 2.6 | < 0.05 |
|  | CHR-C*Visit | 1.2 | 0.73 | 1166.19 | 1.64 | 0.35 |
| A-CPT Q3A Mem | Visit | -1.08 | 1.14 | 1240.56 | -0.95 | 0.55 |
|  | CHR-NCNR*Visit | 1.72 | 0.85 | 519.81 | 2.03 | 0.15 |
|  | CHR-R*Visit | 1.86 | 0.86 | 476.34 | 2.16 | 0.13 |
|  | CHR-C*Visit | 4.97 | 1.55 | 867.68 | 3.2 | < 0.01 |
| A-CPT Q3 INT | Visit | 2.31 | 1.62 | 1219.19 | 1.43 | 0.27 |
|  | CHR-NCNR*Visit | -0.33 | 1.11 | 502.53 | -0.3 | 0.82 |
|  | CHR-R*Visit | -0.69 | 1.1 | 466.77 | -0.62 | 0.67 |
|  | CHR-C*Visit | -0.54 | 1.97 | 825.46 | -0.27 | 0.82 |
| CPT IP | Visit | -0.18 | 0.05 | 1136.73 | -3.33 | < 0.01 |
|  | CHR-NCNR*Visit | 0.04 | 0.04 | 456.47 | 0.99 | 0.45 |
|  | CHR-R*Visit | 0.06 | 0.04 | 418.85 | 1.66 | 0.2 |
|  | CHR-C*Visit | -0.03 | 0.06 | 770.93 | -0.42 | 0.77 |
| NAB Mazes | Visit | 0.13 | 0.36 | 1217.36 | 0.37 | 0.88 |
|  | CHR-NCNR*Visit | -0.23 | 0.24 | 478.37 | -0.99 | 0.71 |
|  | CHR-R*Visit | 0.3 | 0.24 | 443.74 | 1.28 | 0.69 |
|  | CHR-C*Visit | 0.27 | 0.43 | 806.96 | 0.62 | 0.88 |
| Cat Fluency gen | Visit | -0.93 | 0.47 | 1296.92 | -1.96 | 0.24 |
|  | CHR-NCNR*Visit | -0.08 | 0.32 | 510.96 | -0.25 | 0.9 |
|  | CHR-R*Visit | 0.07 | 0.32 | 478.6 | 0.22 | 0.9 |
|  | CHR-C*Visit | -0.31 | 0.59 | 834.34 | -0.52 | 0.79 |
| Cat Fluency cor | Visit | -0.99 | 0.49 | 1430.22 | -2.01 | 0.26 |
|  | CHR-NCNR*Visit | 0.04 | 0.34 | 944.47 | 0.11 | 0.91 |
|  | CHR-R*Visit | 0.17 | 0.34 | 964.57 | 0.51 | 0.81 |
|  | CHR-C*Visit | -0.09 | 0.62 | 1003.56 | -0.15 | 0.91 |
| WMS Total | Visit | -0.61 | 0.27 | 1262.55 | -2.26 | 0.05 |
|  | CHR-NCNR*Visit | -0.06 | 0.18 | 483.2 | -0.34 | 0.76 |
|  | CHR-R*Visit | -0.06 | 0.19 | 442.16 | -0.3 | 0.76 |
|  | CHR-C*Visit | -0.13 | 0.33 | 812.88 | -0.39 | 0.76 |
| LNS | Visit | -0.71 | 0.25 | 1166.76 | -2.84 | < 0.05 |
|  | CHR-NCNR*Visit | -0.2 | 0.16 | 464.12 | -1.19 | 0.51 |
|  | CHR-R*Visit | 0.26 | 0.17 | 437.26 | 1.57 | 0.4 |
|  | CHR-C*Visit | 0.08 | 0.29 | 796.79 | 0.26 | 0.84 |
| BVMT R | Visit | -1.21 | 0.47 | 1275.96 | -2.57 | < 0.05 |
|  | CHR-NCNR*Visit | 0.13 | 0.32 | 483.56 | 0.39 | 0.7 |
|  | CHR-R*Visit | 0.51 | 0.33 | 447.55 | 1.56 | 0.22 |
|  | CHR-C*Visit | 0.52 | 0.58 | 821.74 | 0.88 | 0.5 |
| HVLT R | Visit | -0.32 | 0.37 | 1411.88 | -0.86 | 0.63 |
|  | CHR-NCNR*Visit | -0.29 | 0.25 | 1071.29 | -1.16 | 0.54 |
|  | CHR-R*Visit | -0.15 | 0.26 | 1061.67 | -0.59 | 0.78 |
|  | CHR-C*Visit | 0.11 | 0.46 | 1060.84 | 0.24 | 0.84 |
| BACS-SC | Visit | -1.42 | 0.93 | 1199.33 | -1.53 | 0.43 |
|  | CHR-NCNR*Visit | -0.49 | 0.6 | 471.02 | -0.81 | 0.71 |
|  | CHR-R*Visit | 0.79 | 0.6 | 439.23 | 1.33 | 0.5 |
|  | CHR-C*Visit | 0.75 | 1.07 | 790.87 | 0.7 | 0.72 |
| Trail A | Visit | -0.06 | 0.92 | 1305.59 | -0.07 | 0.97 |
|  | CHR-NCNR*Visit | 0.74 | 0.68 | 612.83 | 1.09 | 0.71 |
|  | CHR-R*Visit | -1.17 | 0.7 | 561.93 | -1.67 | 0.46 |
|  | CHR-C*Visit | -1.22 | 1.17 | 916.66 | -1.04 | 0.71 |
| UPSIT | Visit | -1.73 | 0.39 | 1221.4 | -4.42 | < 0.001 |
|  | CHR-NCNR*Visit | 0.05 | 0.27 | 741.94 | 0.18 | 0.86 |
|  | CHR-R*Visit | -0.1 | 0.27 | 766.99 | -0.39 | 0.76 |
|  | CHR-C*Visit | -0.37 | 0.52 | 859.62 | -0.72 | 0.7 |
| UPSIT BSIT | Visit | -0.9 | 0.16 | 1204.83 | -5.77 | < 0.001 |
|  | CHR-NCNR*Visit | 0.01 | 0.12 | 713.42 | 0.08 | 0.94 |
|  | CHR-R*Visit | 0.03 | 0.11 | 745.21 | 0.24 | 0.94 |
|  | CHR-C*Visit | -0.29 | 0.22 | 884.71 | -1.32 | 0.51 |
| Babble Total | Visit | 2.62 | 1.05 | 1227.46 | 2.51 | 0.15 |
|  | CHR-NCNR*Visit | 0.26 | 0.75 | 513.66 | 0.35 | 0.99 |
|  | CHR-R*Visit | 0.17 | 0.75 | 465.92 | 0.23 | 0.99 |
|  | CHR-C*Visit | -0.07 | 1.3 | 822.65 | -0.05 | 0.99 |
| Babble Longest | Visit | -0.2 | 0.17 | 1511.34 | -1.14 | 0.87 |
|  | CHR-NCNR*Visit | 0.12 | 0.13 | 1250.64 | 0.91 | 0.89 |
|  | CHR-R*Visit | 0.19 | 0.14 | 1209.24 | 1.41 | 0.87 |
|  | CHR-C*Visit | -0.19 | 0.25 | 1195.29 | -0.76 | 0.89 |

*Adjusted for Age at Neuropsych appointment, income, sex, years of education, and mother’s education. SE: Standard Estimate, df: Degrees of Freedom, T: LMM t statistic, q: corrected p-value, HC: Healthy Controls, CHR-NCNR: Clinical High Risk Non-Converter Non-Remitter, CHR-R: Clinical High Risk - Remitter; CHR-C: Clinical High Risk - Converter;* WRAT-4: *Wide Range Achievement Test, 4th Edition – Reading*, WASI IQ: *WASI-II Full Scale IQ (2-subtest version)*, WASI BD: *WASI-II Block Design,* WASI Vocab: *WASI-II Vocabulary*, A-CPT QA Vigil: *Auditory Continuous Performance Test, QA Vigilance*, A-CPT Q3A Mem: *Auditory Continuous Performance Test, Q3A Memory*, A-CPT Q3 INT: *Auditory Continuous Performance Test, Q3 Interference*, CPT-IP: *Continuous Performance Test – Identical Pairs*, NAB Mazes: *Neuropsychological Assessment Battery – Mazes,* Category Fluency gen/cor: *Category Fluency – number of words generated / correct words,* WMS Total: *Wechsler Memory Scale – Total Score*, LNS: *University of Maryland Letter-Number Sequencing*, BVMT R: *Brief Visuospatial Memory Test - Revised,* HVLT R: *Hopkins Verbal Learning Test - Revised*, BACS-SC: *Brief Assessment of Cognition in Schizophrenia – Symbol Coding*, Trail A: *Trail Making Test: Part A*, UPSIT/UPSIT BSIT: *University of Pennsylvania Smell Identification Test*, Babble Total: *Total Word Count in Babble Task,* Babble Longest*: Longest Continuous Phrase in Babble Task.*

#### Clinical Characteristics and Medication

At baseline, there were significant differences between the groups on all four scales for prodromal symptoms scores (p < 0.001), where HCs scored significantly lower on all the sub-scores, followed by higher scores in the CHR-R. CHR-C had the highest scores, followed by CHR-NCNR and then CHR-R. CHR-R had the highest rates of antipsychotic medication usage at baseline (10.2%), followed by CHR-C (5.6%), CHR-NCNR (3.9%), and HC (0%) (Table 6). Over time, CHR-C’s positive symptoms become significantly worse. While both CHR-NCNR and CHR-R improve on all SOPS subcategories, the CHR-R improvement is much greater (Fig 5, Table 7).

**Figure 5:**
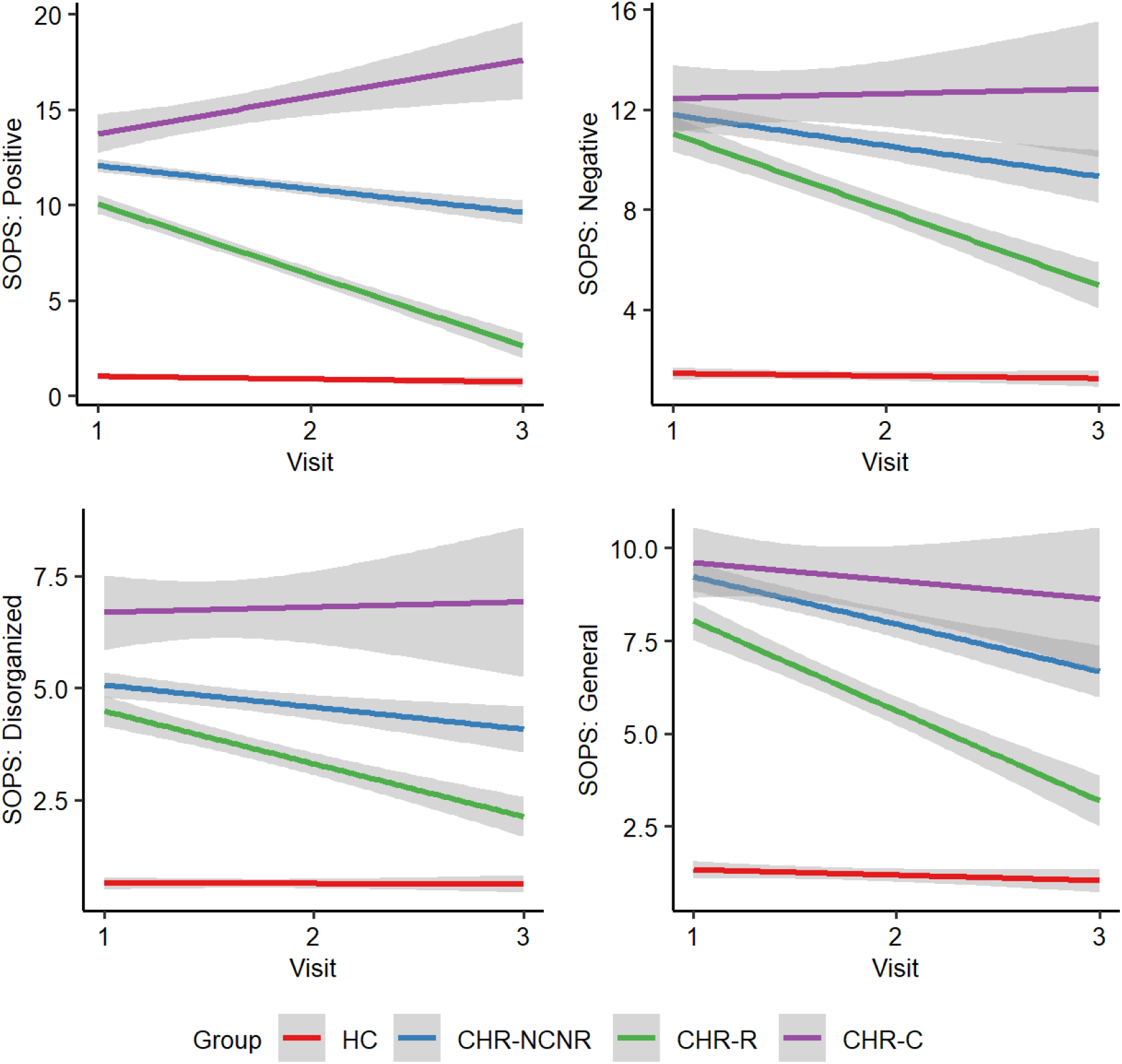
Longitudinal SOPS scores between groups HC: Healthy Controls, CHR-NCNR: Clinical High Risk Non-Converter Non-Remitter, CHR-R: Clinical High Risk - Remitter; CHR-C: Clinical High Risk - Converter; SOPS: Scale of Prodromal Symptoms; Visit 1: Baseline, Visit 2: 12-month or converter visit after baseline, Visit 3: 24-month or converter visit after Visit 2.

**Table 6:** Clinical Symptoms and medication status at baseline between groups.

|  | HC (N=265) | CHR-NCNR(N=384) | CHR-R (N=216) | CHR-C (N=89) | DF | *X*<sup>2</sup>/ H | p | ηp<sup>2</sup>/V |
|---|---|---|---|---|---|---|---|---|
| SIPS: Positive | 1.057 (1.674) | 12.214 (3.508) | 10.657 (4.192) | 13.477 (3.705) | 3 | 698.8 | <0.001 | 0.688 |
| SIPS: Negative | 1.487 (2.277) | 11.974 (6.066) | 11.428 (5.938) | 12.640 (6.448) | 3 | 246.7 | <0.001 | 0.439 |
| SIPS: General | 1.299 (2.123) | 9.390 (4.112) | 8.547 (4.423) | 9.742 (4.371) | 3 | 279.1 | <0.001 | 0.47 |
| SIPS Disorganized | 0.645 (1.178) | 5.160 (3.027) | 4.736 (2.952) | 6.596 (3.982) | 3 | 186.6 | <0.001 | 0.371 |
| Medication status at time of baseline neuropsych |  |  |  |  | 6 | 146.9 | <0.001 | 0.273 |
| Antipsychotic | 0 (0%)* | 15 (3.9%) | 22 (10.2%) | 5 (5.6%) |  |  |  |  |
| None | 265 (100%) | 369 (96.1%) | 194 (89.8%) | 84 (94.4%) |  |  |  |  |

*Note: +: Kruskal-Wallis Test, HC: Healthy Controls, CHR-NCNR: Clinical High Risk Non-Converter Non-Remitter, CHR-R: Clinical High Risk - Remitter; CHR-C: Clinical High Risk - Converter; SOPS: Scale of Prodromal Symptoms; \*: critical value +/- 1.96, DF: Degrees of Freedom, X<sup>2</sup>/H: Pearson’s X<sup>2</sup> test statistic for categorical variables / Kruskal-Wallis H statistic for continuous variables, p: p-value, ηp<sup>2</sup>: Partial Eta Squared; V: Cramers-V.*

**Table 7:**
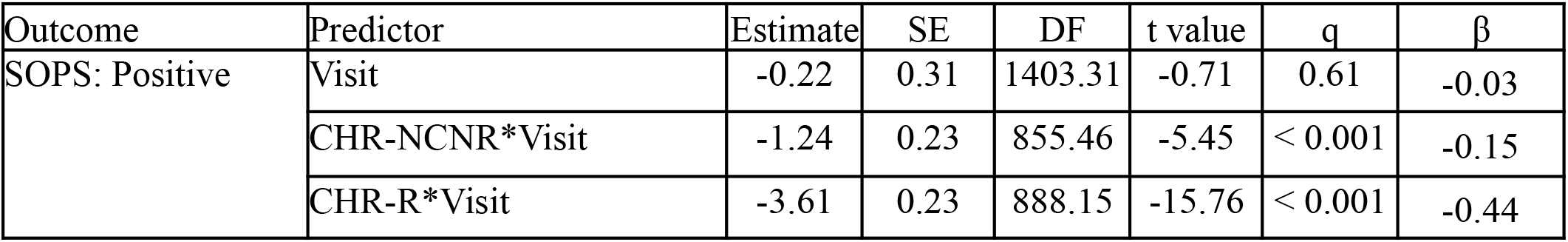
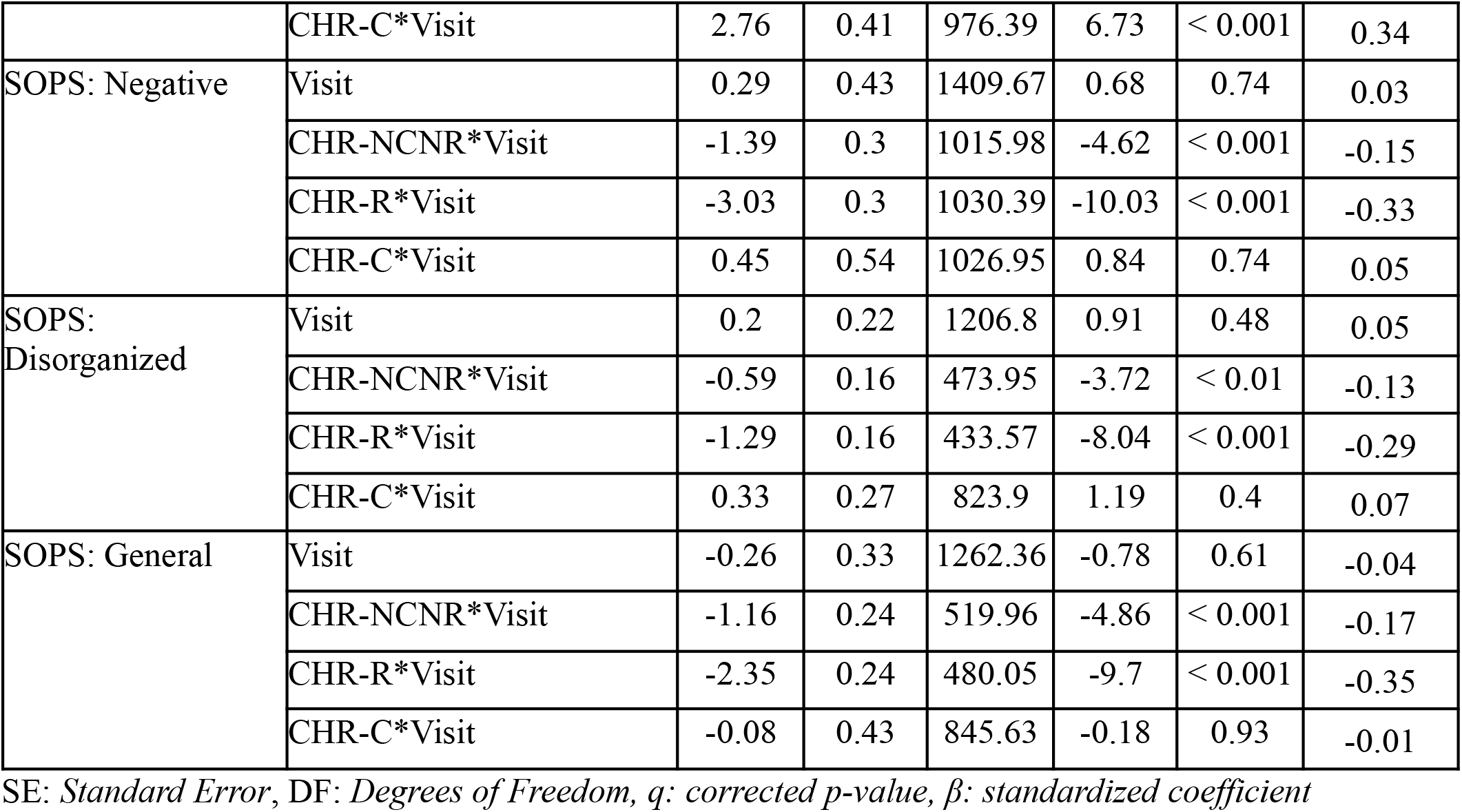
Linear mixed effect models assessing longitudinal clinical outcomes across non converters/non remitters, remitters, and converters.

| Outcome | Predictor | Estimate | SE | DF | t value | q | β |
|---|---|---|---|---|---|---|---|
| SOPS: Positive | Visit | -0.22 | 0.31 | 1403.31 | -0.71 | 0.61 | -0.03 |
|  | CHR-NCNR*Visit | -1.24 | 0.23 | 855.46 | -5.45 | < 0.001 | -0.15 |
|  | CHR-R*Visit | -3.61 | 0.23 | 888.15 | -15.76 | < 0.001 | -0.44 |
|  | CHR-C*Visit | 2.76 | 0.41 | 976.39 | 6.73 | < 0.001 | 0.34 |
| SOPS: Negative | Visit | 0.29 | 0.43 | 1409.67 | 0.68 | 0.74 | 0.03 |
|  | CHR-NCNR*Visit | -1.39 | 0.3 | 1015.98 | -4.62 | < 0.001 | -0.15 |
|  | CHR-R*Visit | -3.03 | 0.3 | 1030.39 | -10.03 | < 0.001 | -0.33 |
|  | CHR-C*Visit | 0.45 | 0.54 | 1026.95 | 0.84 | 0.74 | 0.05 |
| SOPS: Disorganized | Visit | 0.2 | 0.22 | 1206.8 | 0.91 | 0.48 | 0.05 |
|  | CHR-NCNR*Visit | -0.59 | 0.16 | 473.95 | -3.72 | < 0.01 | -0.13 |
|  | CHR-R*Visit | -1.29 | 0.16 | 433.57 | -8.04 | < 0.001 | -0.29 |
|  | CHR-C*Visit | 0.33 | 0.27 | 823.9 | 1.19 | 0.4 | 0.07 |
| SOPS: General | Visit | -0.26 | 0.33 | 1262.36 | -0.78 | 0.61 | -0.04 |
|  | CHR-NCNR*Visit | -1.16 | 0.24 | 519.96 | -4.86 | < 0.001 | -0.17 |
|  | CHR-R*Visit | -2.35 | 0.24 | 480.05 | -9.7 | < 0.001 | -0.35 |
|  | CHR-C*Visit | -0.08 | 0.43 | 845.63 | -0.18 | 0.93 | -0.01 |

SE: *Standard Error*, DF: *Degrees of Freedom, q: corrected p-value, β: standardized coefficient*

## Discussion

To our knowledge, this longitudinal investigation of cognition in CHR is the largest completed study to date to include conversion, remission, and persistent symptoms (i.e., CHR-NCNR) in the same model. Our findings affirm our previous baseline analyses with the NAPLS-2 sample showing that baseline cognitive measures in CHR often differ significantly from control participants and each other, and are selectively in alignment with clinical outcomes (4). The longitudinal analyses show that performance on multiple cognitive measures improved from baseline values over the follow-up period, likely showing an amenability to practice effects to varying extents in different groups.

The fact that cognitive differences are present at baseline and remain significantly different despite adjusting for age, socioeconomic status, and education supports the notion that these differences are core vulnerability markers rather than secondary consequences of illness progression. Some cognitive measures were relatively spared, however, and these are also informative. Specifically, WRAT 4 Reading performance suggested at least average premorbid intellectual ability, though it is possible that modest declines in academic achievement or intellectual ability occurred earlier in life (15). By contrast, the impairments in learning, attention, working memory and processing speed mirror those of classical schizophrenia-related cognitive deficits more closely (2,38). These relatively intact cognitive measures rely on crystallized abilities, visuospatial construction, and perceptual or sensorimotor processes that can be resilient to early developmental disruptions (39–41).

The differences between the CHR subgroups are especially important. Markers of conversion risk, which are present before psychosis onset in our study, include semantic retrieval, executive control, processing speed, and visual scanning, as well as learning deficits, which reinforce the prior evidence about their presence as a neurodevelopmental vulnerability rather than an established psychosis symptoms-induced decline. This shows that conversion is associated with greater severity of cognitive dysfunction within a population that already demonstrates cognitive decline, which highlights the importance of evaluating not just general cognitive impairment but how much impairment.

It is notable that CHR-NCNR outperformed CHR-R on the Babble Test (total words measure). This could mean that, as individuals with CHR remit, they might no longer require certain compensational mechanisms, such as attentional strategies that are required to perceive ‘voices’ in the Babble Test (42). Moreover, it may also serve as a reminder that remission is not equivalent to complete cognitive restoration. The findings that CHR-R do not always outperform CHR-C emphasize this point further, but also suggest that these cognitive deficits are not just an epiphenomenon secondary to symptom severity. Their amenability to improve is a promising indicator of potential responsiveness to therapeutic interventions. The lack of additional cognitive decline post conversion for some variables showing low baseline values (e.g., learning) suggests the possibility of a floor effect, consistent with low baseline performances. Improvements observed postconversion are potentially due to test familiarity, though improvement in remitters on crystalized cognitive domains challenges the idea that this resulted solely from practice effects.

The SOPS scores support the construct validity of the groups, and suggest that the clinical and cognitive vulnerability are associated. Antipsychotic use did not seem to have an observable impact on cognition, since despite greater use in the converters, they remained cognitively impaired while worsening clinically over time. Since CHR-C shows some cognitive improvement despite clinical decline, and CHR-R demonstrates cognitive improvement with symptom amelioration indicates that cognition cannot be interpreted broadly to align with clinical measures, but that specific cognitive domains align with specific clinical measures.

While we are not aware of previous categorizations of cognition in CHR into trait- and state-like variables, the current findings are consistent with recent studies showing relationships between high risk for psychosis states and relatively poorer performance on tests of learning and memory (43). Although most studies have focused on verbal tests of memory in CHR, our findings emphasize the importance of demonstrating deficits in both verbal and visual tests of memory. This finding strengthens the view that declarative learning and memory are significantly involved, rather than only part of a cognitive domain involving verbal processing, as might be suspected, for example, from performance difficulties in verbal learning and memory considered along with difficulties in tests of verbal fluency.

Deficits in declarative memory are among the most consistent and the largest cognitive vulnerabilities in schizophrenia (44,45). While deficits in visual declarative memory are not as well documented as those in verbal declarative memory, they appear to be similar in nature in genetic and clinical high risk participants (4,46). A recent report by Zhang et al (47) showed that both verbal (HVLT-R) and visual (BVMT-R) tests of learning and memory, along with a test of visual working memory (WMS Spatial Span), were related prominently to nonremission based on cognitive trajectories from baseline to a 2-month follow-up. These and other studies have reported associations between lower declarative memory performance and poorer clinical status (47). Lower CHR performance in verbal memory also predicted more rapid conversions in the NAPLS-1 sample than cognitive performances in other domains (26), while Lee et al (48) reported that higher immediate and delayed recall (and other cognitive domains) at baseline were associated with higher rates of remission. Recently, Hedges et al (26) reported significantly lower baseline scores on the Rey Auditory Verbal Learning Test in a CHR group compared to a healthy control group for both immediate and delayed conditions. The immediate recall baseline score was a significant predictor of non-remission and global disability at a two-year follow-up. A recent latent class analysis in a CHR cohort that was followed for 12 months (28) showed a range of performances on a verbal learning and memory test that included an unimpaired subgroup, a group with a stable mild deficit, and a group with an initially larger deficit that ‘caught up’ to the mildly impaired group over the 12-month follow-up period. The stable deficit in the memory test relative to the unimpaired subgroup is of particular note, as it underscores the trait-like nature of the deficit, at least for some individuals.

The trait-like pattern shown by the current findings also aligns well with studies showing that deficits in verbal and visual declarative memory are evident in nonpsychotic relatives of individuals with schizophrenia (46) and are associated with structural abnormalities in brain regions related to processing declarative memory (49–52). Moreover, the low baseline declarative memory scores findings accord well with meta-analyses showing low preprodromal IQ scores in individuals who later developed a schizophrenia-related diagnosis, including the finding that low IQ did not show a relative decline prior to the onset of schizophrenia (53, 54). These studies demonstrate that at least some cognitive abilities, including declarative learning and memory, likely show stable, trait-like deficits before they develop a psychosis-risk syndrome. Although declarative memory deficits appear to cohere well with a trait-state distinction in this study, it does not in any way rule out similar distinctions for other critical cognitive vulnerabilities in our data, such as processing speed or working memory. In fact, findings such as these raise the possibility that state-trait distinctions in cognition might offer a useful way to stratify cognitive performance in CHR and reduce heterogeneity.

### Limitations

The study has a number of limitations. First, like any large longitudinal design, attrition reduced the sample size, which limited power and could have introduced bias. Second, although the data were captured at a consistent time frame, the timing of data capture during conversion naturally varied since it occurred following changes in CHR status. Third, the participants can have a change in clinical status, and not all of them remained in the same status throughout the study. Fourth, practice effects likely contributed to participant improvements, although comparison with HCs mitigates this limitation to some extent. Fifth, antipsychotic and other medications may have influenced our findings, though such effects would likely have been modest.

## Conclusion

Cognition in CHR is associated with multiple longitudinal trajectories that include stable deficits or partial improvements over time, particularly in the remission and on-remission/non-conversion groups. Cognitive improvement in remission, however, is typically not associated with levels associated with full recovery. Improvements tend to involve multiple, though not necessarily all, cognitive domains, while conversion to psychosis tends to include stable deficits in cognition and a narrower band of cognitive reductions. These findings suggest that cognitive trajectories can help distinguish clinical outcomes and reduce heterogeneity in CHR research and treatment strategies.

## Data Availability

NAPLS-2 data are available upon reasonable request, while NAPLS-3 data are available online at the National Institute of Mental Health Data Archive.

https://nda.nih.gov/

## Acknowledgements

First and foremost, we are thankful to the participants in this study. This work was supported by the National Institute of Mental Health (R21MH133001, WY; U01MH081984, JA; R01MH60720, U01MH081944 and K24MH76191, KSC; P50MH066286, CB; U01MH082004, DP; U01MH081988, EW; U01MH082022, SW/KSC; U01MH081902, TC/CB; U01MH076989, DM), made possible by the taxpayers of the United States. We gratefully acknowledge the support of the American people through their investment in scientific research.

## Notes

### Competing Interest Statement

Diana O. Perkins consultants for Alkermes.

### Author Declarations

Ethics committees/IRB of all sites including Emory University, Harvard University, University of Calgary, University of California, Los Angeles, University of California San Francisco, University of California San Diego, University of North Carolina at Chapel Hill, Yale University, and Zucker Hillside Hospital gave ethical approval for this work.

