## Supplementary materials for "Cognitive Trajectories and Clinical Outcomes in the North American Psychosis-Proneness Longitudinal Study (NAPLS2)"

**Table ST1:** Sensitivity analysis comparing effects of converter status variable on cognitive outcomes at baseline when accounting for use of antipsychotic medication at baseline.

| Outcome | Model with No Med | | Model with Med | | P-value comparison |
|---|---|---|---|---|---|
| | Effect sizes and significance values of converter status | | | | |
| | $\eta p^2$ | p | $\eta p^2$ | p | |
| WASI IQ | 0.002 | 0.416 | 0.002 | 0.415 | No change |
| WASI BD | 0.005 | 0.222 | 0.005 | 0.211 | No change |
| WASI Vocab | 0.001 | 0.912 | 0.001 | 0.877 | No change |
| A-CPT Q3 INT | 0.007 | 0.096 | 0.007 | 0.104 | No change |
| CPT IP | 0.001 | 0.549 | 0.001 | 0.660 | No change |
| Cat Fluency gen | 0.015 | 0.007 | 0.015 | 0.005 | No change |
| Cat Fluency cor | 0.016 | 0.007 | 0.016 | 0.005 | No change |
| LNS | 0.010 | 0.053 | 0.010 | 0.058 | No change |
| WMS Total | 0.002 | 0.652 | 0.002 | 0.625 | No change |
| BVMT R | 0.023 | 0.001 | 0.023 | 0.001 | No change |
| HVLT R | 0.011 | 0.031 | 0.011 | 0.029 | No change |
| WRAT 4 | 0.001 | 0.937 | 0.001 | 0.903 | No change |
| A-CPT Q3A Mem | 0.011 | 0.044 | 0.011 | 0.043 | No change |
| NAB Mazes | 0.004 | 0.302 | 0.004 | 0.286 | No change |
| BACS-SC | 0.012 | 0.041 | 0.012 | 0.040 | No change |
| Trail A | 0.014 | 0.012 | 0.014 | 0.011 | No change |
| UPSIT BSIT | 0.002 | 0.436 | 0.002 | 0.537 | No change |
| A-CPT QA Vigil | NA | 0.176 | NA | 0.179 | No change |
| UPSIT | NA | 0.547 | NA | 0.537 | No change |

| | | | | | |
|---|---|---|---|---|---|
| Babble Total | NA | 0.008 | NA | 0.010 | No change |
| Babble Longest | NA | 0.004 | NA | 0.005 | No change |

*Note: ηp²: Partial Eta Squared;* WRAT-4: *Wide Range Achievement Test, 4th Edition – Reading*, WASI IQ: *WASI-II Full Scale IQ (2-subtest version)*, WASI BD: *WASI-II Block Design,* WASI Vocab: *WASI-II Vocabulary*, A-CPT QA Vigil: *Auditory Continuous Performance Test, QA Vigilance*, A-CPT Q3A Mem: *Auditory Continuous Performance Test, Q3A Memory*, A-CPT Q3 INT: *Auditory Continuous Performance Test, Q3 Interference*, CPT-IP: *Continuous Performance Test – Identical Pairs*, NAB Mazes: *Neuropsychological Assessment Battery – Mazes*, Category Fluency gen/cor: *Category Fluency – number of words generated / correct words,* WMS Total: *Wechsler Memory Scale – Total Score*, LNS: *University of Maryland Letter-Number Sequencing*, BVMT R: *Brief Visuospatial Memory Test - Revised,* HVLT R: *Hopkins Verbal Learning Test - Revised*, BACS-SC: *Brief Assessment of Cognition in Schizophrenia – Symbol Coding*, Trail A: *Trail Making Test: Part A*, UPSIT/UPSIT BSIT: *University of Pennsylvania Smell Identification Test*, Babble Total: *Total Word Count in Babble Task,* Babble Longest*: Longest Continuous Phrase in Babble Task.*

**Table ST2:** Linear regression models assessing the association between days since conversion and after conversion cognitive performance while controlling for baseline cognition performance.

| Outcome | Predictors | B | SE | T | P |
|---|---|---|---|---|---|
| WASI BD: After Conversion | WASI BD: Before Conversion | 0.62 | 0.07 | 8.59 | <0.001*** |
| | Days since conversion | -0.01 | 0.01 | -0.52 | 0.6 |
| A-CPT Q3A Mem: After Conversion | CPT.Q3A Mem: Before Conversion | 0.37 | 0.11 | 3.43 | <0.01** |
| | Days since conversion | 0.01 | 0.02 | 0.04 | 0.69 |
| A-CPT Q3 Int: After Conversion | CPT.Q3.INT: Before Conversion | 0.7 | 0.09 | 7.92 | <0.001*** |
| | Days since conversion | -0.02 | 0.02 | -0.87 | 0.39 |
| Trail A: After Conversion | Trail.A: Before Conversion | 0.56 | 0.14 | 4.01 | <0.01** |
| | Days since | -0.01 | 0.01 | -0.55 | 0.59 |

| | |
|---|---|
| | conversion |

WASI BD = *WASI-II Block Design;* A-CPT Q3A Mem = *Auditory Continuous Performance Test, Q3A Memory*; A-CPT Q3 INT = *Auditory Continuous Performance Test, Q3 Interference*; Trail A: Trail Making Test: Part A. Days since conversion is the time between the date of First Episode Psychosis conversion episode and the date of neuropsychological assessment following the conversion episode.

**Table ST3:** Sensitivity analysis comparing effects of converter status variable on cognitive outcomes longitudinally when accounting for use of antipsychotic medication at overtime.

| Outcome | Predictor | Model with No med | | | Model with Med | | | P-value comparison |
|---|---|---|---|---|---|---|---|---|
| | | *b* | SE | p | *b* | SE | p | |
| WRAT-4 | Visit (CHR-NCNR Baseline slope) | -1.22 | 0.43 | < 0.01 | -1.24 | 0.43 | < 0.01 | No change |
| | CHR-R*Visit | -0.02 | 0.19 | 0.91 | -0.02 | 0.19 | 0.89 | No change |
| | CHR-C*Visit | -0.07 | 0.19 | 0.69 | -0.07 | 0.19 | 0.72 | No change |
| WASI IQ | Visit (CHR-NCNR Baseline slope) | 0.88 | 0.94 | 0.35 | 0.96 | 0.95 | 0.32 | No change |
| | CHR-R*Visit | 0.04 | 0.41 | 0.93 | 0.05 | 0.42 | 0.91 | No change |
| | CHR-C*Visit | 0.17 | 0.41 | 0.68 | 0.15 | 0.41 | 0.71 | No change |
| WASI BD | Visit (CHR-NCNR Baseline slope) | 3 | 0.87 | < 0.001 | 3.17 | 0.88 | < 0.001 | No change |
| | CHR-R*Visit | -0.97 | 0.39 | < 0.05 | -0.95 | 0.39 | < 0.05 | No change |
| | CHR-C*Visit | -0.16 | 0.38 | 0.67 | -0.2 | 0.38 | 0.6 | No change |
| WASI Vocab | Visit (CHR-NCNR Baseline slope) | -2.96 | 0.68 | < 0.001 | -2.9 | 0.68 | < 0.001 | No change |
| | CHR-R*Visit | 0.01 | 0.31 | 0.97 | 0.02 | 0.31 | 0.95 | No change |
| | CHR-C*Visit | 0.4 | 0.31 | 0.19 | 0.39 | 0.31 | 0.21 | No change |
| A-CPT QA Vigil | Visit (CHR-NCNR Baseline slope) | 1 | 0.62 | 0.11 | 1 | 0.62 | 0.11 | No change |
| | CHR-R*Visit | -0.08 | 0.33 | 0.82 | -0.08 | 0.33 | 0.82 | No change |
| | CHR-C*Visit | -0.01 | 0.34 | 0.98 | -0.01 | 0.34 | 0.98 | No change |
| A-CPT Q3A Mem | Visit (CHR-NCNR Baseline slope) | 2.8 | 1.32 | < 0.05 | 2.87 | 1.33 | < 0.05 | No change |
| | CHR-R*Visit | -1.05 | 0.68 | 0.12 | -1.04 | 0.68 | 0.13 | No change |
| | CHR-C*Visit | -0.96 | 0.69 | 0.16 | -0.98 | 0.69 | 0.16 | No change |
| A-CPT Q3 INT | Visit (CHR-NCNR Baseline slope) | 1.98 | 1.87 | 0.29 | 2.39 | 1.88 | 0.2 | No change |
| | CHR-R*Visit | 0.17 | 0.88 | 0.85 | 0.24 | 0.88 | 0.79 | No change |
| | CHR-C*Visit | -0.15 | 0.88 | 0.86 | -0.29 | 0.88 | 0.74 | No change |
| CPT IP | Visit (CHR-NCNR Baseline slope) | -0.14 | 0.06 | < 0.05 | -0.11 | 0.06 | 0.07 | **P-value changed.** The slope of CHR-NCNR no longer significantly changed over time, while accounting for effects of medication. |
| | CHR-R*Visit | 0.01 | 0.03 | 0.69 | 0.01 | 0.03 | 0.61 | No change |
| | CHR-C*Visit | 0.04 | 0.03 | 0.2 | 0.03 | 0.03 | 0.34 | No change |
| NAB Mazes | Visit (CHR-NCNR Baseline slope) | 0.68 | 0.41 | 0.1 | 0.73 | 0.41 | 0.08 | No change |
| | CHR-R*Visit | -0.32 | 0.19 | 0.09 | -0.31 | 0.19 | 0.11 | No change |

| | | | | | | | | |
|---|---|---|---|---|---|---|---|---|
| | CHR-C*Visit | 0.2 | 0.19 | 0.28 | 0.19 | 0.19 | 0.32 | No change |
| Cat Fluency gen | Visit (CHR-NCNR Baseline slope) | -0.34 | 0.54 | 0.53 | -0.21 | 0.54 | 0.71 | No change |
| | CHR-R*Visit | 0.08 | 0.26 | 0.74 | 0.11 | 0.26 | 0.67 | No change |
| | CHR-C*Visit | 0.21 | 0.26 | 0.42 | 0.16 | 0.26 | 0.52 | No change |
| Cat Fluency cor | Visit (CHR-NCNR Baseline slope) | -0.18 | 0.56 | 0.74 | -0.03 | 0.56 | 0.95 | No change |
| | CHR-R*Visit | 0.07 | 0.27 | 0.8 | 0.09 | 0.27 | 0.73 | No change |
| | CHR-C*Visit | 0.17 | 0.27 | 0.52 | 0.12 | 0.27 | 0.65 | No change |
| WMS Total | Visit (CHR-NCNR Baseline slope) | -0.79 | 0.31 | < 0.05 | -0.77 | 0.31 | < 0.05 | No change |
| | CHR-R*Visit | 0.01 | 0.15 | 0.97 | 0.01 | 0.15 | 0.95 | No change |
| | CHR-C*Visit | 0.02 | 0.15 | 0.88 | 0.01 | 0.15 | 0.92 | No change |
| LNS | Visit (CHR-NCNR Baseline slope) | -0.57 | 0.28 | < 0.05 | -0.56 | 0.28 | < 0.05 | No change |
| | CHR-R*Visit | -0.24 | 0.13 | 0.06 | -0.24 | 0.13 | 0.07 | No change |
| | CHR-C*Visit | 0.22 | 0.13 | 0.08 | 0.22 | 0.13 | 0.09 | No change |
| BVMT R | Visit (CHR-NCNR Baseline slope) | -0.48 | 0.55 | 0.38 | -0.46 | 0.55 | 0.4 | No change |
| | CHR-R*Visit | -0.24 | 0.26 | 0.37 | -0.23 | 0.26 | 0.37 | No change |
| | CHR-C*Visit | 0.14 | 0.26 | 0.6 | 0.13 | 0.26 | 0.61 | No change |
| HVLT R | Visit (CHR-NCNR Baseline slope) | -0.19 | 0.44 | 0.66 | -0.14 | 0.45 | 0.75 | No change |
| | CHR-R*Visit | -0.17 | 0.21 | 0.41 | -0.16 | 0.21 | 0.44 | No change |
| | CHR-C*Visit | -0.03 | 0.21 | 0.9 | -0.04 | 0.21 | 0.83 | No change |
| BACS-SC | Visit (CHR-NCNR Baseline slope) | -0.88 | 1.03 | 0.4 | -0.6 | 1.04 | 0.56 | No change |
| | CHR-R*Visit | -0.89 | 0.46 | 0.05 | -0.85 | 0.46 | 0.06 | No change |
| | CHR-C*Visit | 0.45 | 0.45 | 0.32 | 0.36 | 0.45 | 0.42 | No change |
| Trail A | Visit (CHR-NCNR Baseline slope) | -1.51 | 1.11 | 0.17 | -1.68 | 1.11 | 0.13 | No change |
| | CHR-R*Visit | 1.23 | 0.58 | < 0.05 | 1.21 | 0.58 | < 0.05 | No change |
| | CHR-C*Visit | -0.66 | 0.58 | 0.25 | -0.61 | 0.58 | 0.3 | No change |
| UPSIT | Visit (CHR-NCNR Baseline slope) | -1.7 | 0.46 | < 0.001 | -1.61 | 0.47 | < 0.001 | No change |
| | CHR-R*Visit | 0.14 | 0.22 | 0.52 | 0.16 | 0.22 | 0.48 | No change |
| | CHR-C*Visit | 0.02 | 0.22 | 0.92 | 0 | 0.22 | 0.99 | No change |
| UPSIT BSIT | Visit (CHR-NCNR Baseline slope) | -1.06 | 0.19 | < 0.001 | -1.04 | 0.19 | < 0.001 | No change |
| | CHR-R*Visit | 0.08 | 0.09 | 0.39 | 0.09 | 0.09 | 0.37 | No change |
| | CHR-C*Visit | 0.1 | 0.09 | 0.26 | 0.1 | 0.09 | 0.28 | No change |
| Babble Total | Visit (CHR-NCNR Baseline slope) | 2.61 | 1.22 | < 0.05 | 2.69 | 1.23 | < 0.05 | No change |

| | | | | | | | | |
|---|---|---|---|---|---|---|---|---|
| | CHR-R*Visit | 0.13 | 0.62 | 0.83 | 0.15 | 0.62 | 0.81 | No change |
| | CHR-C*Visit | 0.02 | 0.62 | 0.97 | -0.01 | 0.62 | 0.99 | No change |
| Babble Longest | Visit (CHR-NCNR Baseline slope) | -0.14 | 0.21 | 0.5 | -0.16 | 0.21 | 0.44 | No change |
| | CHR-R*Visit | 0.08 | 0.11 | 0.46 | 0.08 | 0.11 | 0.49 | No change |
| | CHR-C*Visit | 0.15 | 0.11 | 0.2 | 0.15 | 0.11 | 0.18 | No change |

*Note: b: Beta; SE: Standard Error; CHR-NCNR: Clinical High Risk Non-Converter Non-Remitter, CHR-R: Clinical High Risk - Remitter; CHR-C: Clinical High Risk - Converter;* WRAT-4: *Wide Range Achievement Test, 4th Edition – Reading*, WASI IQ: *WASI-II Full Scale IQ (2-subtest version)*, WASI BD: *WASI-II Block Design,* WASI Vocab: *WASI-II Vocabulary*, A-CPT QA Vigil: *Auditory Continuous Performance Test, QA Vigilance*, A-CPT Q3A Mem: *Auditory Continuous Performance Test, Q3A Memory*, A-CPT Q3 INT: *Auditory Continuous Performance Test, Q3 Interference*, CPT-IP: *Continuous Performance Test – Identical Pairs*, NAB Mazes: *Neuropsychological Assessment Battery – Mazes*, Category Fluency gen/cor: *Category Fluency – number of words generated / correct words,* WMS Total: *Wechsler Memory Scale – Total Score*, LNS: *University of Maryland Letter-Number Sequencing*, BVMT R: *Brief Visuospatial Memory Test - Revised,* HVLT R: *Hopkins Verbal Learning Test - Revised*, BACS-SC: *Brief Assessment of Cognition in Schizophrenia – Symbol Coding*, Trail A: *Trail Making Test: Part A*, UPSIT/UPSIT BSIT: *University of Pennsylvania Smell Identification Test*, Babble Total: *Total Word Count in Babble Task,* Babble Longest*: Longest Continuous Phrase in Babble Task.*

**Table ST4:** Baseline cognitive outcome differences between subjects who dropped out (those who only attended the first visit) and subjects who followed up (those who attended subsequent visits).

| Outcome | Dropped out (N=316) | | Followed-up (N=638) | | D | P |
|---|---|---|---|---|---|---|
| | Mean | SD | Mean | SD | | |
| WASI IQ | 104.04 | 14.86 | 106.55 | 15.50 | -0.16 | 0.11 |
| WASI BD | 45.99 | 14.50 | 46.30 | 15.65 | -0.02 | 0.84 |
| WASI Vocab | 54.61 | 10.41 | 56.58 | 10.23 | -0.19 | 0.06 |
| A-CPT Q3 INT | 56.72 | 20.28 | 57.30 | 21.74 | -0.03 | 0.80 |
| CPT IP | 2.37 | 0.78 | 2.58 | 0.78 | -0.27 | <0.05 |
| Cat Fluency gen | 24.49 | 6.05 | 24.69 | 5.90 | -0.03 | 0.80 |

| | | | | | | |
|---|---|---|---|---|---|---|
| Cat Fluency cor | 23.85 | 6.07 | 24.14 | 5.94 | -0.05 | 0.75 |
| LNS | 14.80 | 3.50 | 15.20 | 3.77 | -0.11 | 0.32 |
| WMS Total | 16.66 | 3.71 | 16.87 | 3.54 | -0.06 | 0.71 |
| BVMT R | 25.39 | 6.13 | 26.21 | 6.06 | -0.13 | 0.23 |
| HVLT R | 25.54 | 5.13 | 26.32 | 4.91 | -0.16 | 0.14 |
| WRAT 4 | 58.31 | 7.43 | 59.05 | 7.16 | -0.10 | 0.38 |
| A-CPT Q3A Mem | 82.02 | 14.30 | 83.37 | 14.03 | -0.10 | 0.41 |
| NAB Mazes | 20.37 | 4.91 | 20.38 | 5.13 | >-0.05 | 0.99 |
| BACS-SC | 57.96 | 11.56 | 59.39 | 14.13 | -0.11 | 0.32 |
| Trail A | 29.08 | 11.71 | 29.10 | 11.59 | -0.05 | 0.99 |
| UPSIT BSIT | 10.17 | 1.51 | 10.05 | 1.59 | 0.08 | 0.59 |
| A-CPT QA Vigil | 95.01 | 8.05 | 95.50 | 7.42 | -0.06 | 0.71 |
| UPSIT | 33.95 | 4.38 | 33.81 | 4.16 | 0.03 | 0.80 |
| Babble Total | 15.22 | 13.02 | 14.61 | 12.36 | 0.05 | 0.75 |
| Babble Longest | 2.94 | 1.72 | 2.87 | 2.18 | 0.03 | 0.80 |

*Note: SD: Standard Deviation; D: Cohen's D; P: FDR-Adjusted P Value;* WRAT-4: *Wide Range Achievement Test, 4th Edition – Reading*, WASI IQ: *WASI-II Full Scale IQ (2-subtest version)*, WASI BD: *WASI-II Block Design,* WASI Vocab: *WASI-II Vocabulary*, A-CPT QA Vigil: *Auditory Continuous Performance Test, QA Vigilance*, A-CPT Q3A Mem: *Auditory Continuous Performance Test, Q3A Memory*, A-CPT Q3 INT: *Auditory Continuous Performance Test, Q3*

*Interference*, CPT-IP: *Continuous Performance Test – Identical Pairs*, NAB Mazes: *Neuropsychological Assessment Battery – Mazes*, Category Fluency gen/cor: *Category Fluency – number of words generated / correct words,* WMS Total: *Wechsler Memory Scale – Total Score*, LNS: *University of Maryland Letter-Number Sequencing*, BVMT R: *Brief Visuospatial Memory Test - Revised,* HVLT R: *Hopkins Verbal Learning Test - Revised*, BACS-SC: *Brief Assessment of Cognition in Schizophrenia – Symbol Coding*, Trail A: *Trail Making Test: Part A*, UPSIT/UPSIT BSIT: *University of Pennsylvania Smell Identification Test*, Babble Total: *Total Word Count in Babble Task,* Babble Longest*: Longest Continuous Phrase in Babble Task*.
